# Proteomic profiling of plasma extracellular vesicles identifies signatures of innate immunity, coagulation, and endothelial activation in septic patients

**DOI:** 10.1101/2025.02.21.25322420

**Authors:** Chanhee Park, Taekyung Ryu, Rashida Mohamed-Hinds, Kyungdo Kim, Jin Hyeok Kim, Lin Zou, Brittney Williams, Chan Hyun Na, Wei Chao

## Abstract

Plasma extracellular vesicles (EVs) are cell-derived lipid particles and reportedly play a role in sepsis pathogenesis. This study aimed to identify EV cargo proteins in septic patients and explore their association with key sepsis pathophysiology. Plasma EVs were subjected to Tandem Mass Tag (TMT)-based quantitative proteomic analysis. We identified 522 differentially expressed (DE) EV proteins in septic patients (n=15) compared to the healthy controls (n=10). The KEGG analysis of the DE proteins revealed multiple functional pathways linked to sepsis, *e.g.,* complement/coagulation, platelet activation, phagosome, inflammation, and neutrophil extracellular trap formation. Weighted Gene Coexpression Network Analysis of 1,642 EV proteins identified nine unique protein modules, some of which were highly correlated with the sepsis diagnosis and diverse plasma markers, including organ injury, inflammation, coagulopathy, and endothelial activation. Cell type-specific enrichment analysis revealed the cellular origins of EVs, including immune and epithelial cells, neurons, and glial cells. Thus, the current study discovered complex proteomic signatures in plasma EVs that are closely associated with key pathophysiological responses in sepsis. These findings support the importance of EV cargo proteins in the patients’ immune responses, coagulation, and endothelial activation and lay the foundation for future mechanistic study of plasma EVs in sepsis pathogenesis.

## Introduction

Sepsis is induced by a dysregulated host response to infection and characterized clinically by hyperinflammation, hemodynamic collapse, endothelial injury, coagulopathy, and multiple organ dysfunction [1,2]. It is the leading cause of in-hospital death in the United States [3,4]. While significant progress has been made in our understanding of sepsis pathophysiology, diagnosis, and intervention, the current management of sepsis is still limited to supportive care without specific target therapy.

Extracellular vesicles (EVs) are cell-derived lipid particles with heterogenous and biologically active cargo molecules containing various proteins and nucleic acids and have emerged as key mediators of intercellular communication under various physiological and pathophysiological conditions [5,6]. Given their multifaceted biological and pathophysiological effects, uncovering the protein composition of EVs could potentially reveal mechanisms of underlying diseases, promote the discovery of novel biomarkers, and refine therapeutic targets [7,8]. In sepsis, EVs have been studied for their roles in pathophysiological responses such as inflammation, coagulation, and various organ dysfunctions [9–11], and as potential biomarkers [9,12]. Identifying EV cargo proteins is a critical first step toward fully understanding the contents and functions of plasma EVs in human sepsis.

In the current study, we conducted mass spectrometry-based proteomic profiling of EV cargo using Tandem Mass Tag (TMT) technology for 15 septic patients admitted to the ICU and 10 healthy participants. The TMT-based proteomic method enables precise proteomic quantification and offers the advantages of high sensitivity, accuracy, and reliability of detection with minimal technical variability [13]. We identify a long list of proteins differentially expressed (DE) in septic EVs. Employing various bioinformatics tools such as the Kyoto Encyclopedia of Genes and Genomes (KEGG) pathway and Gene Ontology (GO), we systematically analyze the functions of various differentially expressed proteins and their specific functional pathways. Using Weighted Gene Co-expression Network Analysis (WGCNA), we also investigate the module-trait relationship between the EV cargo proteins and various traits, such as multiple plasma markers linked to sepsis pathophysiology and pathogenesis as well as clinical laboratory and outcome data. The revelation of the functional pathways associated with the EV proteins offers a mechanistic insight into sepsis pathogenesis.

## Results

### Patient cohort

All patients met the enrollment criteria for sepsis based on suspected or confirmed infection with a concomitant increase in SOFA score ≥ 2. A cohort of 15 septic patients with sepsis in had a mean age of 53.7 (23-78) years and 53% of them were male. Similarly, the healthy controls had a mean age of 40.4 (25-66) years with 25% being male. The average SOFA score for the 15 septic patients at admission was 11.3 (range 6-18) and the 28-day mortality rate was 26.7%. A detailed summary of the demographic and clinical laboratory data can be found in **Table 1**.

**Table 1.** Demographic and clinical data of septic and healthy subjects.

|  | **Healthy (n = 10)** | **Sepsis (n = 15)** |
| --- | --- | --- |
| **Age (years)** | 40.4 (25-66) | 53.7 (23-78) |
| **Sex (M/F)** | 2/8 | 8/7 |
| **Ethnicity** |  |  |
| Black | 2 (20%) | 4 (26.7%) |
| Asian | 0 (0%) | 1 (6.7%) |
| Hispanic | 0 (0%) | 0 (0%) |
| White | 8 (80%) | 10 (66.7%) |
| Other | 0 (0%) | 0 (0%) |
| **SOFA score** |  | 11.3 (6-18) |
| **ICU stay (Day)** |  | 18.1 (4-54) |
| **28-day Mortality** |  | 4 (26.7%) |
| **Hematology Labs** |  |  |
| INR ($\leq 1.2$) | | 1.6 (1.1-3) |
| PTT (30-40 sec) |  | 37.2 (23-66) |
| Platelets (150-300 $\times 10^3/\mu\text{L}$) | | 195.7 (41-445) |
| **Chemistry Labs** |  |  |
| Lactate ($<2$ mmol/L) | | 2.8 (1-11.5) |
| Creatinine (0.7-1.3 mg/dL) |  | 1.7 (0.38-3.34) |

Demographics with average (range) or count (percent).
INR, international normalized ratio; PTT, partial thromboplastin time.

### Plasma EV characterization

As illustrated in **Figure 1**, we isolated plasma EVs from 10 healthy control (HC) individuals and 15 septic patients using ultracentrifugation. We first optimized the EV purification steps by increasing the numbers of washing steps and ultracentrifugation cycles using control plasma samples. Three rounds of washing and ultracentrifugation enabled us to identify the highest percentage of exosome proteins, according to the comparison with the ExoCarta Top 100 list, enriching exosome proteins to 7.33% with the 409 total identified proteins **(Supplemental Table S1**). Thus, we used the three times of ultracentrifugation and washing to enrich EVs using plasma samples from sepsis patients and HC individuals. After the EV enrichment, we conducted a nanoparticle tracking analysis (NTA) to evaluate the density and size of the isolated EV samples. The numbers of plasma EVs were 5.8±3.8 x 10^9^/mL and 16.6±11.3 x 10^9^ /mL (mean ± SD) in the HC individuals and septic patients, respectively (**Figure 2A**). The number of septic EVs showed a significant increase compared to HC EVs, with *P* value of 0.0031. The average sizes of the isolated EVs were 98.9 ± 9.4 nm in HC individuals and 100.2 ± 10.1 nm in septic patients (**Figure 2B**), with overall even size distributions between the sepsis patients and HC individuals (**Figure 2C)**. Additionally, we examined the relative abundance of CD9 and Alix (PDCD6IP), markers of endosomal EVs by Western blot **(Figure 2D)**. Alix is reportedly crucial for exosome formation and secretion, while CD9 is a tetraspanin protein expressed on the exosome surface. [14] Across all samples from HC individuals and septic patients, Alix and CD9 were found on the expected sizes, and it suggests that EVs were successfully enriched in all samples.

**Figure 1.**
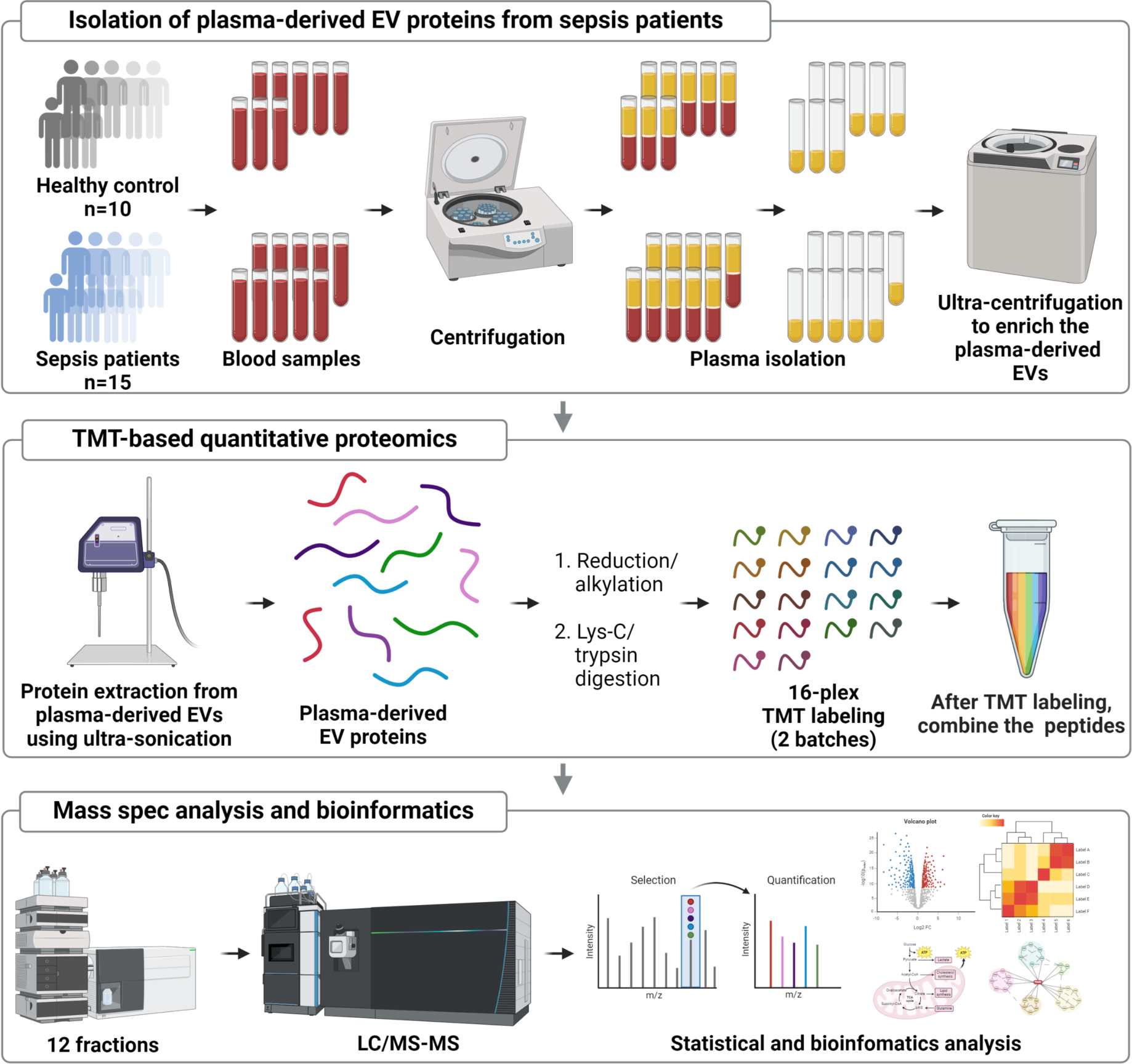
TMT-based quantitative proteomic analysis of plasma EVs in septic and healthy subjects. Human plasma samples were prepared from 15 septic patients and age/sex-matched 10 HC individuals. Plasma extracellular vehicles (EVs) were purified using ultra-centrifugation. Cargo proteins were extracted from EVs using ultra-sonication and subsequently subjected to reduction, alkylation, and enzymatic digestion. The peptides were then labeled with TMT reagents and separated into 24 fractions before being subjected to mass spectrometry and statistical and bioinformatic analyses.

**Figure 2.**
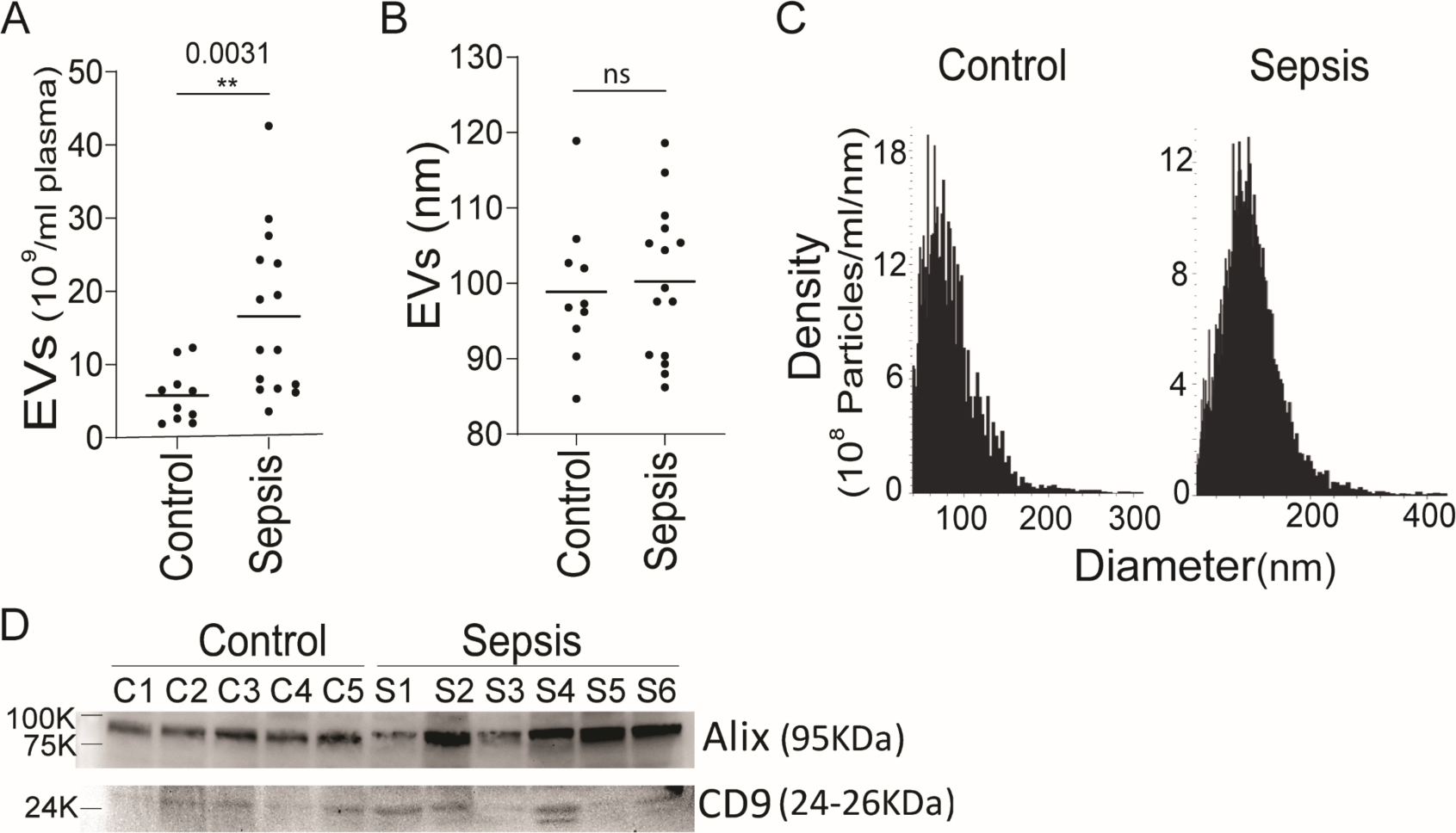
Characterization of plasma EVs from septic patients and health controls. The concentration of EVs (**A**) and the mean EV size (**B**) isolated from the plasma of Healthy control (Control) individuals and septic patients (Sepsis) were quantified using a nanoparticle tracking analysis (Viewsizer 3000, Horiba Scientific). Comparative analysis was conducted based on EV samples from 15 septic patients and 10 HC individuals. The bar in the middle of the dots indicates the mean value. An unpaired t-test was used for the statistical analysis (**: *P* ≤ 0.01, and ns: not significant). (**C**) The representative size distribution for two EV samples is presented. (**D**) Western blot of Alix and CD9 in the EVs isolated from a group of HC individuals and septic patients. Alix: PDCD6IP (Programmed cell death 6-interacting protein), CD9: tetra-spanin membrane protein.

### Proteomic profiling of EV cargo proteins

To profile plasma EV cargo proteins, we conducted the TMT-based quantitative proteomic analysis of the EV proteins. We acquired 1,918,398 MS/MS spectra with 154,249 spectra matched to peptides. This resulted in the identification of 18,348 peptides and 2,371 proteins. A total of 1,642 proteins were quantified across all 25 samples without any missing values (**Supplemental Table S2 and S3**). The statistical analysis revealed 522 proteins that were differentially expressed with a q-value < 0.05, which represents a False Discovery Rate (FDR), in septic patients as compared to healthy individuals (**Figure 3A)**. Out of the 522 differentially expressed proteins, 301 proteins were upregulated, which included neutrophil defensin 1 and 3 (DEFA1, DEFA3), ADP-ribosylation factor 5 (ARF5), nicotinamide phosphoribosyl transferase (NAMPT), histone H4 (H4C1), mitogen-activated protein kinase 4 (MAPK4), C-reactive protein (CRP), serum amyloid A-1 protein (SAA1), histone HIST2H3PS2 (H3-2), histone H2A type 2-C (H2AC2). Among the 221 downregulated EV proteins from the lowest q-value were CD5 antigen-like (CD5L) proteins, haptoglobin-related protein (HPR), Immunoglobulin heavy constant mu (IGHM), Immunoglobulin heavy variable 2-5 (IGHV2-5), Immunoglobulin heavy variable 3/OR16-13 (IGHV3OR16-13), Immunoglobulin lambda variable 3-27 (IGLV3-27), and apolipoprotein L1 (APOL1) **(Supplemental Table S4**). To evaluate the discrimination capability of the differentially expressed proteins between the sepsis and HC groups, we conducted principal component analysis (PCA) of the 522 differentially expressed EV proteins, which revealed a clear separation between the septic patients and healthy controls with no overlap (**Figure 3B**). To determine functional pathways associated with the differentially expressed proteins, we conducted KEGG pathway analysis and identified the multiple enriched pathways that include the complement and coagulation cascades as the most enriched pathway with the lowest *P* value (56.08 of −Log_10_ (*P* value)), followed by phagosome pathway, malaria infection-related pathways, neutrophil extracellular trap formation, platelet activation pathway and a number of other pathways **(Figure 3C, Supplemental Table S5). Supplemental Figure S1** illustrates 30 out of 138 proteins in the complement and coagulation systems, which were the most enriched according to the KEGG analysis and their functionally associated pathways. To investigate further the key proteins in the enriched pathways of the differential EV proteins, we conducted protein-protein interaction (PPI) analyses using the STRING database **(Supplemental Figure S2A∼2D)**. Among the proteins enriched in the complement and coagulation cascades pathway, proteins such as Fibrinogen alpha chain (FGA), Vitronectin (VTN), and Plasma protease C1 inhibitor (SERPING1) were the key proteins **(Supplemental Figure S2A)**. Among the proteins enriched in the phagosome pathway, which is most enriched after the complement and coagulation cascades, Platelet glycoprotein 4 (CD36), Integrin alpha-M (ITGAM), and Integrin beta-2 (ITGB2) were shown as the key proteins **(Supplemental Figure S2B)**. In the Malaria pathway, which implies a response to an infection, ICAM1, CD36, and Platelet endothelial cell adhesion molecule (PECAM1) were among the important proteins **(Supplemental Figure S2C)**. In the neutrophil extracellular trap formation pathway, ICAM1, ITGB2, and Low-affinity immunoglobulin gamma Fc region receptor II-a (FCGR2A) were among the important proteins **(Supplemental Figure S2D)**. Subsequently, we conducted a Gene Ontology Cellular Components (GO-CC) analysis to determine the subcellular origins of the differentially expressed proteins in the EVs **(Figure 3D and Supplement Table S6)**. The extracellular space was the most enriched cellular compartment, followed by extracellular region, extracellular exosome, extracellular vesicle, extracellular organelle, and extracellular membrane-bound organelle in the analysis. These GO-CC results suggest that the differentially expressed EV proteins in septic patients are primarily derived from extracellular compartments, confirming their origin within the EV proteome and highlighting the extracellular space and vesicle-related compartments as major sources.

**Figure 3.**
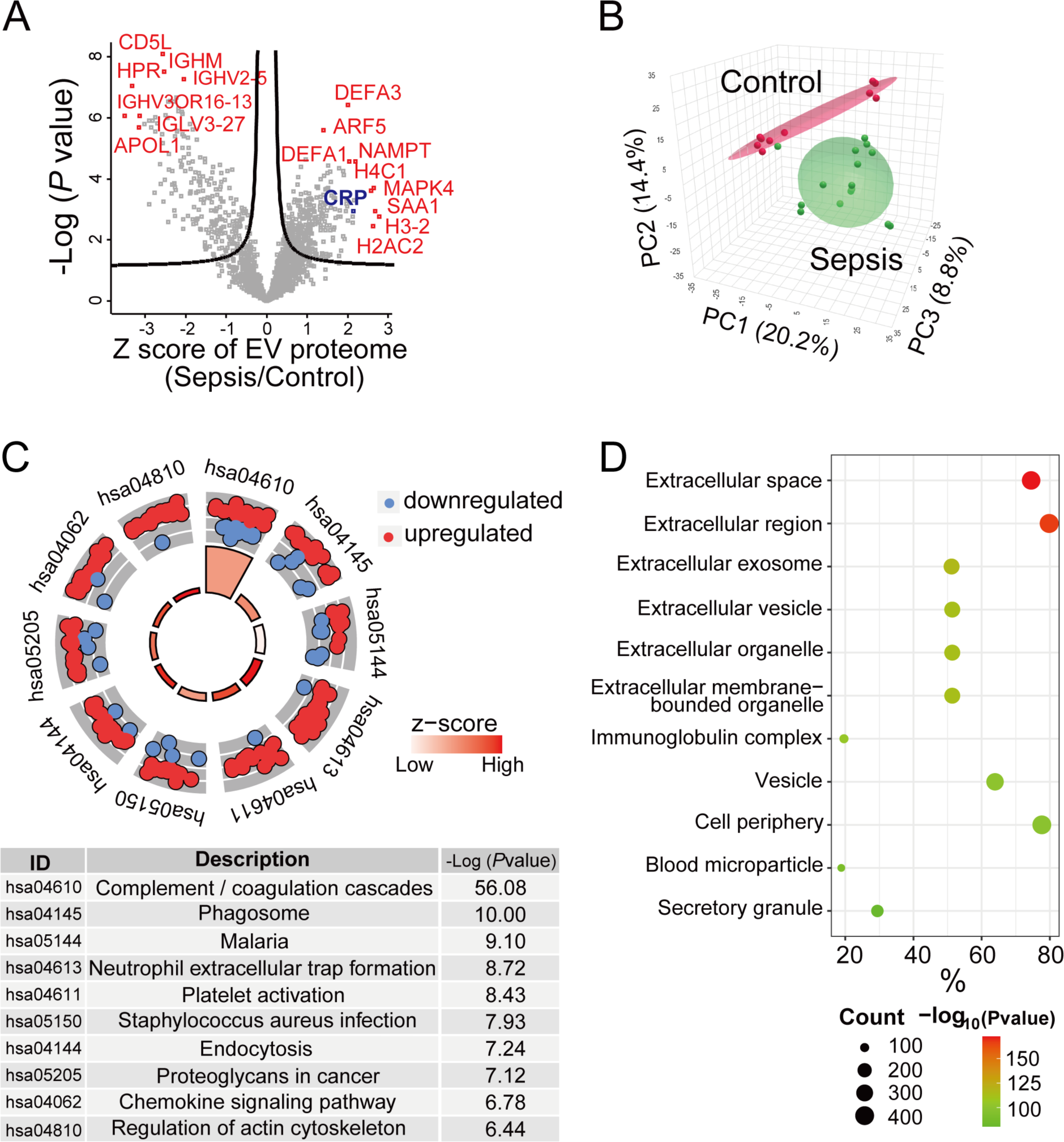
Proteomic analysis of differentially expressed EV proteins in septic patients compared to HC individuals. (**A**) A volcano plot for the plasma EV proteins from 15 septic patients and 10 HC individuals. The x-axis represents the fold-change of septic patients/HC individuals in the Z score scale (a standardized measure that indicates how far a data point is away from the mean of a distribution. Z score of 1 presents 1 standard deviation). The y-axis represents the *P* value of statistical analysis in −Log_10_ scale. The curved lines indicate the boundary for a q-value of 0.05. Proteins with q < 0.05 are differentially expressed in septic patients, and representative proteins among them are highlighted in red. (**B**) Principal Component Analysis (PCA) for the 522 differentially expressed EV proteins between septic (n=15) and HC individuals (n=10). (**C**) The enriched pathways in the KEGG Pathway analysis were displayed using the GO plot package. The GO plot shows the up/down-regulation of differentially expressed proteins in sepsis, displaying the −Log_10_ (*P* value) and z-score values for each pathway. The red circles represent upregulated proteins in each pathway, while the blue circles represent downregulated proteins. The size of the trapezoids in the inner circle of the GO circle reflects the −Log_10_ (*P* value) of the enriched pathway. The color of the trapezoids corresponds to the z-score. The z-score in the package is calculated as follows: z-score = (the number of upregulated proteins – the number of downregulated proteins)/square root (the number of proteins). (**D**) GO analysis of Cellular Components (GO-CC) describes where the differentially expressed EV proteins originate from. GO-CC enrichment analysis selected the top 11 as representative lists based on −Log_10_ (*P* value). The count represents the number of enriched proteins in GO terms. The % represents the proportion of genes in the input list associated with the specific GO term.

### EV protein clusters are closely correlated with sepsis diagnosis

To further understand the potential role of the EV cargo proteins in sepsis pathophysiology, we conducted WGCNA with the total EV proteins that have been quantified – with or without differential expression – and obtained protein clusters (modules) that have similar expression patterns across the plasma samples **(Supplemental Figures S3-S4)**, representing a group of proteins potentially from the same pathways [15]. The WGCNA yielded 9 modules (**Supplemental Table S7**). Among them, the M3, M4, and M6 modules exhibited the highest correlation with the sepsis diagnosis (*P* value ≤ 0.001) followed by M5 and M7 with 0.01 < *P* ≤ 0.05 **(Figure 4A, Supplemental Table S8)**. Interestingly, M3 and M4 modules were negatively correlated with the sepsis diagnosis trait, showing down-regulation of the eigenprotein (meta-expression profile of the proteins in the module) in sepsis (**Figures 4B and 4D)**, while the M5, M6, and M7 modules were positively correlated with the sepsis diagnosis, showing up-regulation of eigenprotein (**Figures 4F, 4H, and 4J).** To find key proteins, we examined module membership (MM) and protein significance (PS) values in each module. For the M3 module, Haptoglobin-related protein (HPR), Hepatocyte growth factor activator (HGFAC), and CD5L were the key proteins, exhibiting high correlation with sepsis diagnosis and strongly representing the M3 module. **(Figure 4C)**. These proteins are considered as key proteins involved in the sepsis pathogenesis process. For the M4 module, Apolipoprotein L1 (APOL1), Apolipoprotein A1 (APOA1), and Myeloid and Erythroid Nuclear Termination Stage-Specific Protein (MENT) were the key proteins, exhibiting high correlation with sepsis diagnosis and strongly representing the M4 module. **(Figure 4E)**. For the M5 module, H4C1, MAPK4, and Histone 3.1 (H3C1) are the key proteins, exhibiting high correlation with sepsis diagnosis and strongly representing the M5 module. **(Figure 4G)**. For the M6 module, Defensin A1 (DEFA1), High-affinity immunoglobulin epsilon receptor subunit gamma (FCER1G), and Leukosialin (SPN) were the key proteins, exhibiting high correlation with sepsis diagnosis and strongly representing the M6 module **(Figure 4I)**. Similarly, in **the** M7 module, CRP, Lipopolysaccharide-binding protein (LBP), and Ceruloplasmin (CP) were the key proteins, exhibiting a high correlation with sepsis diagnosis and strongly representing the M7 module. **(Figure 4K)**.

**Figure 4.**
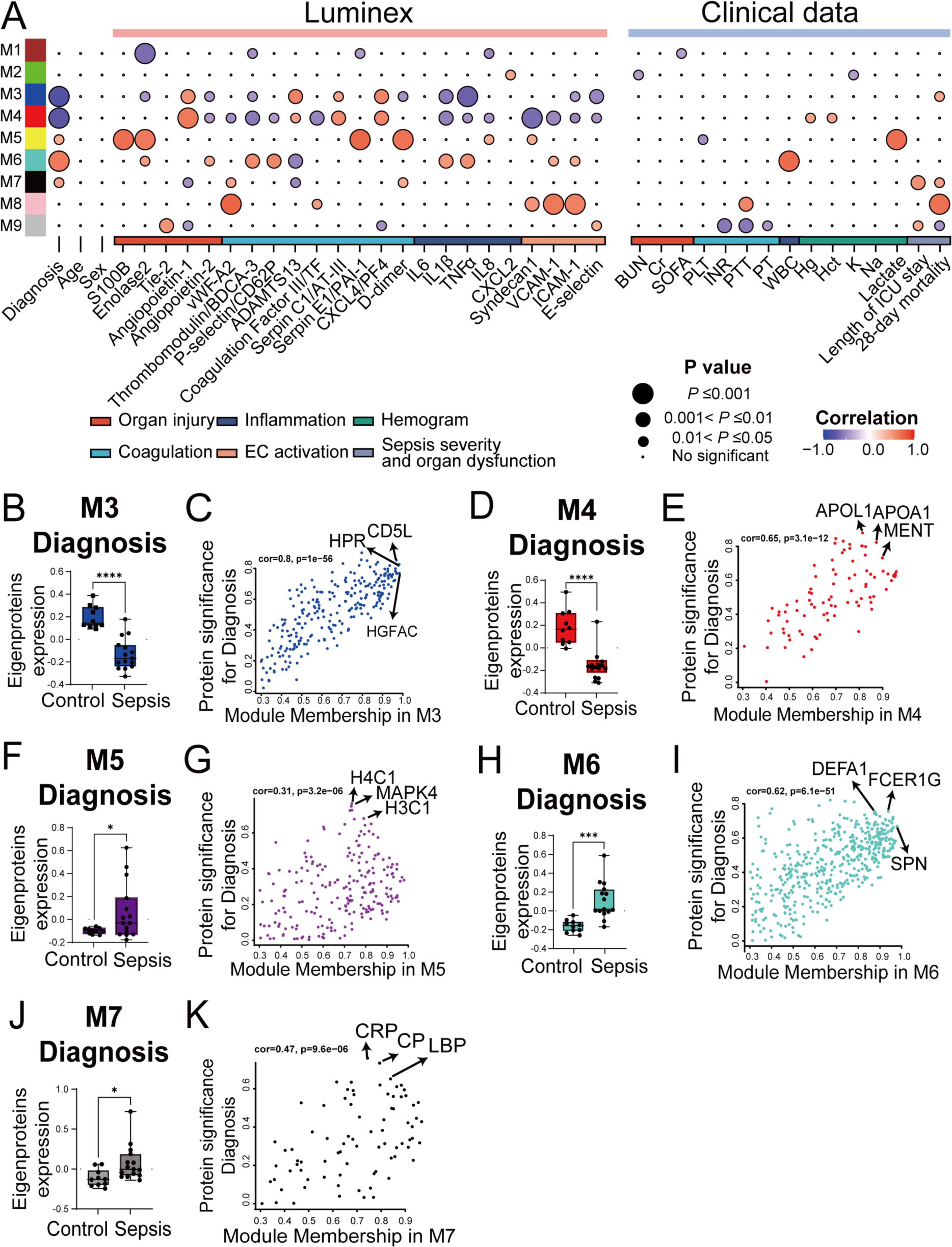
The module–trait relationships by the WGCNA analysis of plasma-derived EV proteome data. (**A**) A heatmap illustrating Pearson correlations between eigenprotein expression levels of 9 WGCNA modules and the values of the sepsis-related traits of the plasma samples. The meanings of abbreviations for the traits in clinical lab data are as follows: BUN: blood urea nitrogen, Cr: creatinine, SOFA: sequential organ failure assessment, PLT: platelet count, INR: international normalized ratio, PTT: partial thromboplastin, PT: prothrombin time, WBC: White blood cell count, Hg: hemoglobin, Hct: hematocrit, K: potassium, and Na: sodium. The correlations were color-coded on a scale ranging from 1 (indicating a positive correlation, red) to −1 (indicating a negative correlation, blue). The size of each circle corresponds to the *P* value, while the color indicates the correlation value. (**B, D, F, H, J**) Relative eigenprotein abundance for the modules which showed a significant correlation with the diagnosis between the sepsis and healthy control group was shown on the box plot; M3 (B), M4 (D), M5 (F), M6 (H), M7 (J). (**C, E, G, I, K**) MM-PS plots show the relationship between the module membership (MM) for each module and protein significance (PS) for sepsis diagnosis; M3 (C), M4 (E), M5 (G), M6 (I), and M7 (K). The key driving proteins were marked by black arrows in the plots. An unpaired t-test was used for the statistical analysis of eigenprotein expression (*, *P* < 0.05; ***, *P* < 0.001; ****, *P* < 0.0001).

### EV protein clusters are associated with pathophysiological markers of sepsis

The pathophysiology of sepsis is complex and heterogeneous from patient to patient. To capture this, we designed a set of plasma markers known to be linked to sepsis pathophysiology, including organ injury, coagulation, inflammation, and endothelial cell (EC) activation. As illustrated in **Figure 4A**, using a Luminex multiplex platform and clinical laboratory tests, we measured the plasma samples from the two groups of participants for their organ injury markers (S100B, enolase2, Tie-2, angiopoietin-1, angiopoietin-2, BUN, Cr, SOFA), coagulation markers (vWF-A2, thrombomodulin/BDCA-3, P-selectin/CD62P, ADAMTS13, coagulation factor III/tissue factor, serpin C1/AT-III, serpin E1/PAI-1, CXCL4/PF4, D-dimer, Platelet, INR, PTT, PT), inflammation markers (IL6, IL1β, TNFα, IL8, CXCL2, WBC), EC activation markers (syndecan1, VCAM-1, ICAM-1, E-selectin), hemogram (Hg, Hct, Hg, K, Na, Lactate), and the clinical outcomes (length of ICU stay, 28-day mortality).

We discovered that all the plasma markers in the Luminex panel, except for CXCL2 and Tie2 (Angiopoietin-1 receptor), were markedly upregulated in the septic cohort compared with the healthy controls (**Supplemental Table S9**). Moreover, as shown in **Figure 4A**, the modules highly related to sepsis diagnosis, such as M3, M4, M5, M6, and M7, were significantly correlated to sepsis-related pathophysiological markers. Among the markers, M3 module were highly correlated with Angiopoietin-1, ADAMTS13, CXCL4/PF4, IL1β, TNFα, and E-selectin, whereas M4 module was closely correlated with angiopoietin-1, thrombomodulin, Coagulation factor/tissue factor, serpin C1/AT-III, CXCL4/PF4, IL1β, syndecan-1, and VCAM-1. The correlated traits with the M5 module were S100B, Enolase2, Serpin E1/PAI-1, and D-dimer. For M6 module, thrombomodulin/BDCA-3, P-selectin/CD62P, ADAMTS13, IL1β, and TNFα are significantly regulated. In M7 module, there were no traits correlated with *P* ≤ 0.01. Interestingly, in the right panel of **Figure 4A**, the WBC traits exhibited a high correlation with the M6 module, which, not surprisingly, may suggest that M6’s correlation with the coagulation and inflammation traits are probably linked to circulating WBC or vice versa. Another noticeable result in WGCNA was that the M8 module showed a high correlation with 28-day mortality even it was not correlated to sepsis diagnosis. Since the M8 module is highly correlated with the traits related to coagulation (vWF, coagulation factor III, and PTT) and EC activation (syndecan1, VCAM1, and ICAM1), it suggests that these traits may be connected to the 28-day mortality in these septic patients.

**Figure 5** shows the top player proteins in each of these EV protein modules that showed a close correlation with the physiological traits of sepsis. Specifically, we plotted PS and MM of each protein in the modules to determine significant proteins from M3 to M6 modules in relation to their respective top 4 traits beside sepsis diagnosis trait with significant correlation with *P* ≤ 0.01 (**Figures 5** and **Supplemental Table S8**). Other traits that exhibited significance at *P* < 0.01 in the M3 (A), M4 (B), and M6 (C) modules, but not presented in **Figure 5**, are displayed in **Supplemental Figure S5**.

**Figure 5.**
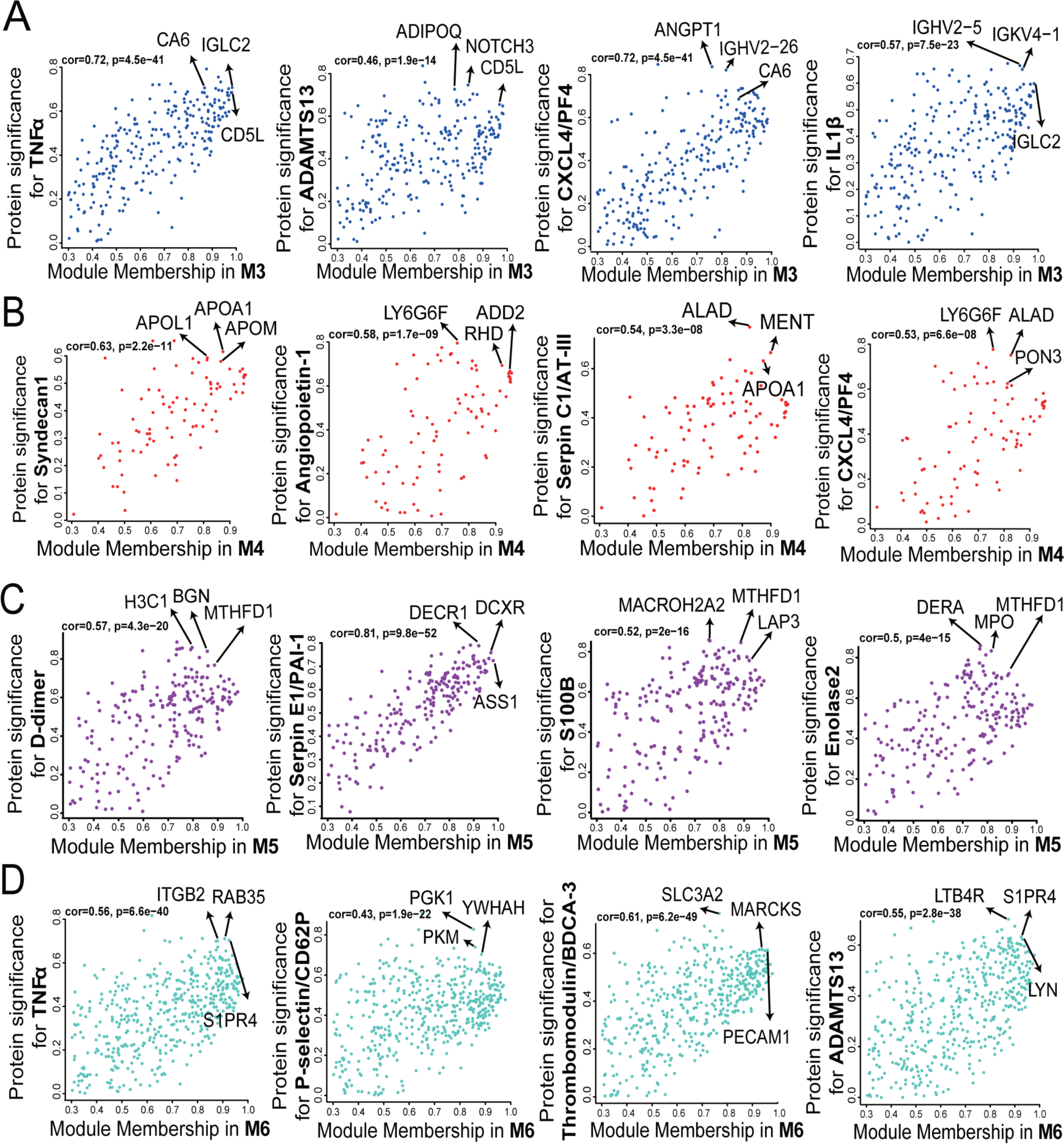
The Module membership (MM) and protein significance (PS) plots in the M3-M6 modules. The traits in Luminex that correlate to M3 ∼ M6 modules in *P* ≤ 0.01 were selected to analyze MM-PS plots and find significant proteins in sepsis pathophysiology. (A) MM-PS plots show the relationship between module membership (MM) for the M3 module and protein significance (PS) for the top 4 traits except diagnosis (TNFα, ADAMTS13, CXCL4/PFA4, IL1β). The key driving proteins were marked by black arrows. The protein list in the plot is presented in **Supplemental Table S8**. (B) MM-PS plots show the relationship between MM for the M4 module and PS for the top 4 traits except diagnosis (Syndecan1, Angiopoietin-1, Serpin C1/AT-III, CXCL4/PF4). (C) MM-PS plots show the relationship between MM for the M5 module and PS for the top 4 traits except diagnosis (D-dimer, SERPINE1, S100B, and Enolase2). (D) MM-PS plots show the relationship between MM for the M6 module and PS for the top 4 traits except diagnosis (TNFα, P-selectin/CD62P, thrombomodulin/BDCA3, ADAMTS13).

### Cell-type and pathway analyses of the WGCNA module proteins

Since the M3, M4, and M6 modules were identified as critical modules showing strong correlations with the sepsis diagnosis (*P* ≤ 0.001), and to a lesser degree with M5 and M7 modules (0.001 < *P* ≤ 0.01), each of the five modules was subjected to cell-type as well as KEGG pathway enrichment analysis to identify the specific cell types from which the proteins are originated and the functional pathways they belong to. We utilized the DISCO database, which contains information on cell-type marker genes annotated through single-cell RNA sequencing, and the top 10 enriched cell types were selected based on −Log_10_ (*P* value) **(Figure 6)**. The M3 proteins were enriched mainly for epithelial cells, immune cells, glial cells, and fibroblasts **(Figure 6A)**. The most enriched pathways for these proteins were the complement and coagulation cascades by KEGG pathway analysis (**Figure 6B, Supplemental Table S11**). Since the M3 proteins were mostly downregulated in sepsis, these data suggest that the M3 proteins related to the pathways were likely downregulated during sepsis, such as coagulation factor V (F5) in the coagulation cascade and C4b-binding protein (C4BP) in the complement cascade **(Supplement Figure S1 and Supplement Table S4)**. The M4 proteins were enriched for fibroblasts, immune cells, epithelial cells, and neuronal cells, and functionally linked to the proteasome pathway (**Figure 6C-6D)**. The M5 module was enriched for immune cells and mainly related functionally to proteasome and metabolic pathways **(Figure 6E-6F)**. The M6 proteins were enriched for hematopoietic stem cells and arterial endothelial cells and functionally related to actin cytoskeleton, leukocyte trans-endothelial migration, and platelet activation (**Figure 6G-6H)**. The M7 module was enriched for neuronal cells, melanocytes, epithelial cells, and fibroblasts (**Figure 6I)**. The complement and coagulation cascades were the most enriched pathway for the M7 module (**Figure 6J)**. The module proteins are related to the upregulation of the pathways, such as CD59 in the complement cascade and coagulation factor X (F10) in the coagulation cascade **(Supplement Figure S1 and Supplement Table S4)**. Finally, to access key proteins in each related pathway, we conducted interactome analysis for the modules using STRING PPI **(Supplemental Figure S6-S10, Supplemental Table S10)**. While C1Q family proteins related to the complement and coagulation cascades are noticeable in the M3 module, the apolipoprotein family in the M4 module was examined significantly. In the M5 module, proteins related to metabolic pathways, such as the Elongation factor family (EEF), Alcohol dehydrogenase 6 (ADH6), and histone proteins, were key proteins. In the M6 module, Cell division control protein 42 homolog (CDC42) and Profilin-1 (PFN1) in the actin filament organization and proteins of some integrin beta (ITGB) family in the extracellular matrix organization were significantly detected, while in the M7 module, Antithrombin-III (SerpinC1) and Complement factor I (CFI) were key proteins.

**Figure 6.**
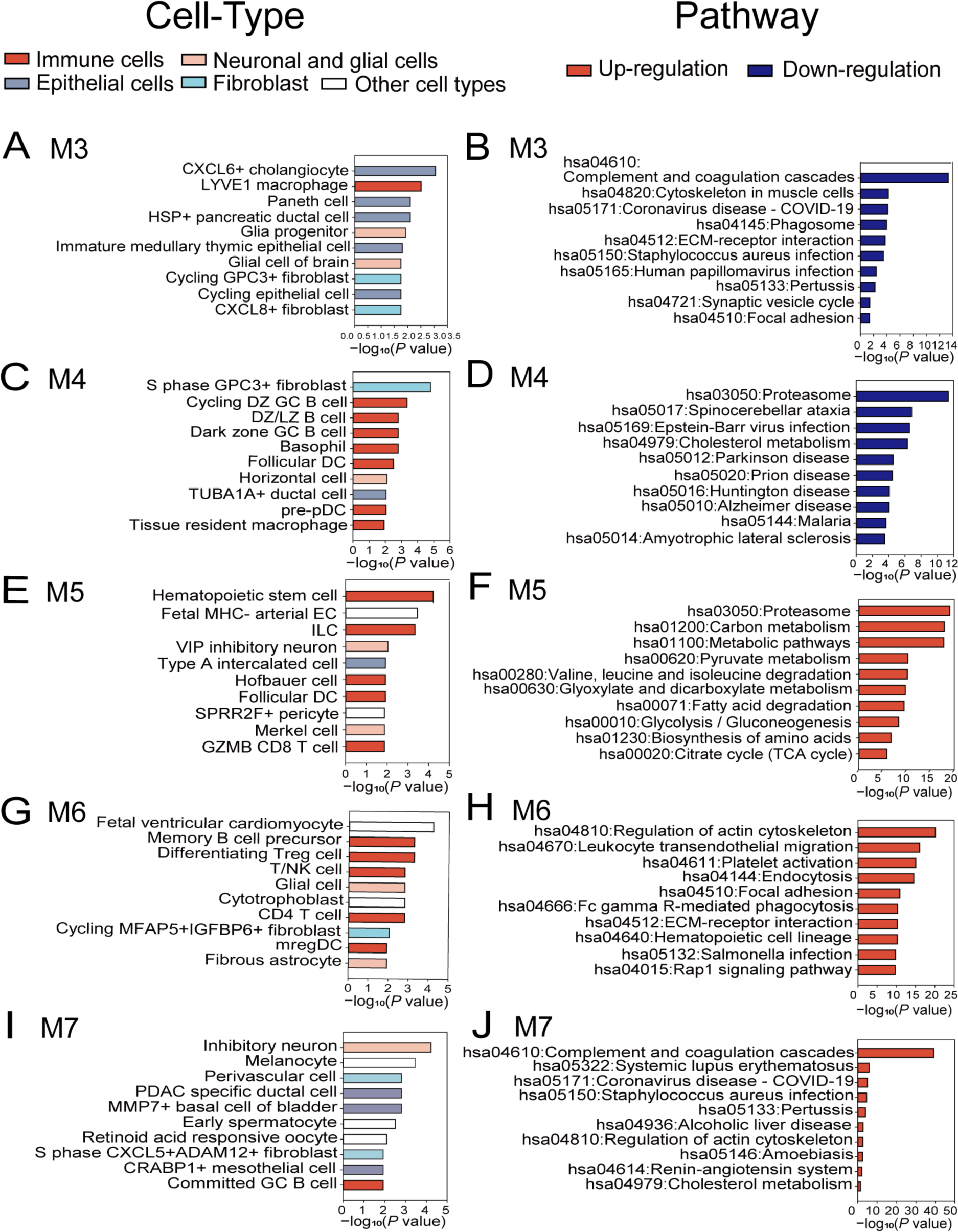
Cell-type enrichment and pathway analysis of the EV proteins in the M3, M4, M5, M6, and M7 modules. (A, C, E, G, I) Cell-type enrichment analysis for proteins in the modules using the DISCO database, highlighting the top 10 enriched cell types for each module, based on −Log_10_ (*P* value). Each bar color represents a specific cell type. (A) is for M3, (C) for M4, (E) for M5, (G) for M6, and (I) for M7. (B, D, F, H, J) KEGG pathway analysis for the modules was performed and the top 10 significant pathway terms were selected for each module based on *P* value, showing upregulated pathways in red and downregulated pathways in blue. (B) is for M3, (D) is for M4, (F) is for M5, (H) is for M6, and (J) is for M7.

### Selection of EV proteins highly correlated with the sepsis diagnosis

Finally, to test the discrimination abilities of EV candidate markers for sepsis, we performed ROC analyses on the 522 differentially expressed proteins in the sepsis cohort (**Supplemental Table S12)**. We selected the top highly discriminative proteins in AUCs > 0.8 with a significant Z score as shown in **Table 2**. With the Z score, we narrowed down the proteins more correlated with the sepsis diagnosis among many strong AUC proteins. The candidates, all closely associated with sepsis diagnosis, are grouped in the four functional pathways - complement and coagulation cascade, immune responses, endothelial activation and metabolic pathways. Integrin beta-2 (ITGB2), von Willebrand factor (VWF), and Inter-alpha-trypsin inhibitor heavy chain H4 (ITIH4) are the top three of the upregulated markers in the complement and coagulation pathways. ADP-ribosylation factor 5 (ARF5), Golgi-associated plant pathogenesis-related protein 1 (GLIPR2), and Sphingosine 1-phosphate receptor 4 (S1PR4) are the top three of them in the immune responses, while Neutrophil defensin 1 (DEFA1), DFEA3, and S100-A9 (Protein S100-A9) are the top three in the endothelial activation. H4C1 (Histone H4), MAPK4 (Mitogen-activated protein kinase 4), and SLC16A3 (Monocarboxylate transporter 4) are the top three in the metabolic pathways. While further validation is needed, these proteins seemed highly correlated to sepsis diagnosis.

**Table 2.** ROC analysis of selected EV cargo proteins correlated highly with sepsis diagnosis.

| Gene name | Gene description | AUC | Z score | Reference |
| --- | --- | --- | --- | --- |
| ***Complement and Coagulation Cascade*** |  |  |  |  |
| *Upregulation* |  |  |  |  |
| ITGB2 | Integrin beta-2 | 0.96 | 1.44 | [19] |
| VWF | von Willebrand factor | 0.95 | 1.96 | [20] |
| ITIH4 | Inter-alpha-trypsin inhibitor heavy chain H4 | 0.94 | 1.39 | [24] |
| SERPINE1 | Plasminogen activator inhibitor 1 (PAI-1) | 0.91 | 1.32 | [59] |
| SAA1 | Serum amyloid A-1 protein | 0.93 | 2.68 | [60] |
| SERPINA1 | Alpha-1-antitrypsin | 0.86 | 1.92 | [34] |
| *Downregulation* |  |  |  |  |
| PF4 | Platelet factor 4 | 0.97 | -2.77 | [61] |
| ***Immune responses*** |  |  |  |  |
| *Upregulation* |  |  |  |  |
| ARF5 | ADP-ribosylation factor 5 | 1 | 1.40 | [25] |
| GLIPR2 | Golgi-associated plant pathogenesis-related protein 1 | 1 | 1.79 | [26] |
| S1PR4 | Sphingosine 1-phosphate receptor 4 | 0.99 | 1.45 | [27] |
| SPN | Leukosialin | 0.97 | 1.58 | [29] |
| CXCR2 | C-X-C chemokine receptor type 2 | 0.92 | 1.65 | [62] |
| *Downregulation* |  |  |  |  |
| HGFAC | Hepatocyte growth factor activator | 1 | -2.09 | [30] |
| HPR | Haptoglobin-related protein | 0.98 | - 3.32 | [31] |
| ***Endothelial activation*** |  |  |  |  |
| *Upregulation* |  |  |  |  |
| DEFA1 | Neutrophil defensin 1 | 0.99 | 2.04 | [32] |
| DEFA3 | Neutrophil defensin 3 | 0.99 | 2.01 | [32] |
| S100A9 | Protein S100-A9 | 0.99 | 1.52 | [33] |
| S100A8 | Protein S100-A8 | 0.96 | 1.56 | [33] |
| CDC42 | Cell division control protein 42 homolog | 0.86 | 0.81 | [35] |
| *Downregulation* |  |  |  |  |
| TNN | Tenascin-N | 0.97 | -2.71 | [63] |
| ***Metabolic pathways*** |  |  |  |  |
| *Upregulation* |  |  |  |  |
| H4C1 | Histone H4 | 0.99 | 2.64 | [40] |
| MAPK4 | Mitogen-activated protein kinase 4 | 0.97 | 2.58 | [36] |
| SLC16A3 | Monocarboxylate transporter 4 | 0.97 | 1.97 | [37] |
| CP | Ceruloplasmin | 0.96 | 1.51 | [38] |
| H3C1 | Histone H3.1 | 0.95 | 2.15 | [40] |
| *Downregulation* |  |  |  |  |
| APOL1 | Apolipoprotein L1 | 0.99 | -3.16 | [41] |

The ROC analysis was performed to estimate the discriminating power between the sepsis and control groups for the selected driving proteins in the M3, M4, M5, M6, and M7 modules overlapped with differentially expressed proteins in sepsis with $q \leq 0.05$ . Among the proteins with $AUC \geq 0.8$ , the top 5 to 6 in the highest Z score (upregulation) or one or two of the lowest Z score (downregulation) were selected, which represent intense discrimination and most fold changes. The values in parentheses indicate the lower and upper AUC values within the 95% confidence interval of the repeated AUC analyses conducted by 500 bootstrappings.

### EV cargo proteome vs. plasma proteome

Although the EV cargo proteins could be a useful resource for the sepsis diagnosis, many of the EV proteins can be detectable from the plasma proteome such as vWF and SERPINE1 (**Supplemental Table S13**). We compared the EV cargo proteome with the plasma proteome from the same subjects. Plasma proteome analysis identified 8,658 peptides (**Supplemental Table S14)** and 1,072 proteins (**Supplemental Table S15)**, 309 of them were differentially expressed in the sepsis patients **(Figure 7A, Supplemental Table S16)**. Among the 1072 proteins, 797 proteins were shared between plasma and EV proteomes, and 275 and 1574 proteins were unique in the plasma and EVs, respectively (**Figure 7B)**. Among the 309 differentially expressed proteins in the plasma proteome, 116 proteins were shared between plasma and EV proteomes, and 193 and 406 proteins were unique in the plasma and EVs, respectively (**Figure 7C)**. Among the 406 differentially expressed proteins unique to EVs, 302 proteins were quantified only in EV, while 104 proteins were quantified in both EV and plasma proteome but differential only in EV proteome. This data suggests that ∼50% (104 out of 220 proteins) of the plasma proteome that overlap with the differential EV proteome were not differential in the plasma proteome, emphasizing the importance of analyzing EV proteome to understand the functional pathways of sepsis. To better understand the correlation between the expression level of EV and plasma proteome, we conducted a correlation analysis between them. The median correlation coefficients between EV and plasma proteome were ∼0.42 for total proteomes **(Figure 7D)** and ∼0.65 for differentially expressed proteins **(Figure 7E)**, indicating a moderate correlation between them. These results suggest that a significant number of potential sepsis-related protiens identified from EVs would not be detectible or not differential in the plasma proteome, emphasizing the importance of EV proteins in understanding sepsis functional pathways.

**Figure 7.**
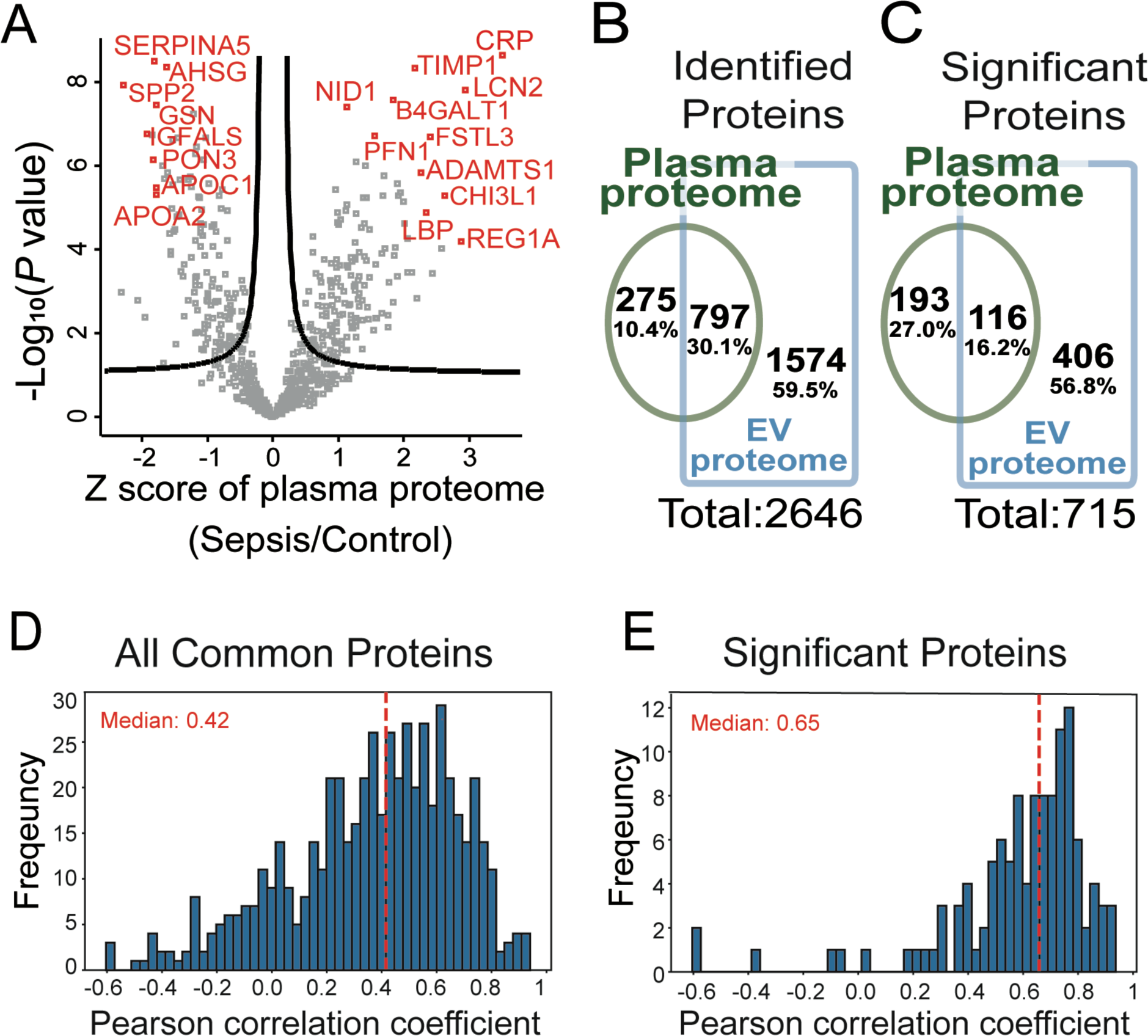
Comparative analysis of differentially expressed proteins between the EV and plasma proteomes in septic patients. (A) The volcano plot displays the differentially expressed proteins between septic patients and HC in the plasma proteome. The x-axis represents the Z score, indicating the magnitude of change, while the y-axis represents the −Log_10_ (*P* value), indicating the statistical significance. The most representative upregulated and downregulated proteins were colored in red. (B) The Venn diagram shows the overlapping proteins between the total plasma proteome and the total EV proteome. (C) The Venn diagram illustrates the overlapping differentially expressed proteins between the plasma proteome and the EV proteome. (D-E) Histograms show the protein abundance correlations between plasma proteins and EV proteins for all quantified proteins (D) and differentially expressed proteins (E) at the level of individual samples. The red dashed line indicates the median correlation coefficient value for each proteome.

## Discussion

We discovered that more than 500 EV cargo proteins were differentially expressed in septic patients when compared to that of healthy controls. Network analysis identified four functional pathways linked to these proteins: complement and coagulation cascades, inflammation and immune responses, endothelial activation and vascular injury, and metabolic pathways. These functional pathways are linked to and have been characterized as an integrated part of the complex sepsis pathophysiology that determines the clinical outcome of sepsis, suggesting the importance of EV cargo proteins in sepsis pathogenesis.

Sepsis is often associated with complement and coagulation dysfunction [16] [17]. Supporting the role of plasma EVs in sepsis pathogenesis of innate immune activation and procoagulation, the bioinformatics analysis with KEGG pathway and STRING PPI revealed the complement and coagulation cascades and platelet activation as the most enriched pathways associated with the EV cargo proteins. Moreover, as detailed in Table 2, several top players in septic EVs associated with complements and coagulation cascades have high AUC values, suggesting their potential role in the sepsis pathophysiology. ITGB2 (CD18), an important integrin expressed on leukocytes, consists of a part of the complement receptor complexes CD11a/CD18 and CD11b/CD18 and binds to ICAM-1 on endothelial cells [18], affecting neutrophil function in sepsis [19]. VWF is reportedly increased in septic patients [20] and contributes to endothelial cell dysfunction [21]. WVF also has been studied as a sepsis biomarker candidate with ADAMTS13 and has been integrated into clinical use in conjugation with other clinical parameters [22]. SERPINE1 or plasminogen activator inhibitor 1 (PAI-1) inhibits plasminogen activation to lead to impaired fibrinolysis, which is important to prevent excessive clotting [23]. ITIH4 (Inter-alpha-trypsin inhibitor heavy chain H4) has not been well studied in sepsis but the upregulation of ITIH4 is detected in LPS-induced systemic inflammation of pigs [24].

We identified a few enriched pathways that are closely related to inflammation and host immune responses, such as phagosome, infection-related pathways, endocytosis, and chemokine signaling pathways. The top proteins with high AUC values are ARF5, GLIPR2, S1PR4, CXCR1, SPN, CXCR2, HGFAC, and HPR. ARF5 is involved in the ADP-ribosylation factor family of guanine-nucleotide-binding (G) proteins and primarily associated with membrane traffic [25]. GLIPR2 plays a role in innate immune regulation related to Toll-like receptor 4 [26]. S1PR4 is abundant in immune cells, suggesting that the increased S1PR4 may be involved in the dysregulated immune response in sepsis [27]. Blocking of CXCR1 and CXCR2 is reported to attenuate the severity of lipopolysaccharide-induced hyperinflammation [28]. SPN is expressed on most leukocytes including T-cells, neutrophils, and monocytes, and promotes inflammation through induction of proinflammatory cytokines such as TNFα, IL12, and IL6 [29]. Among the downregulated EV cargo proteins, HGFAC has not been studied in sepsis, but HGF was studied as an anti-inflammatory regulator in sepsis [30], which indicates a potential role of HGFAC on sepsis-associated inflammation. HPR in septic plasma is reported higher in septic patients than in those only with local infections [31]. In our cohort, the HPR level in plasma was not significantly changed but decreased in EVs. Interestingly, in the M3 module, many immunoglobulin-related proteins were also significantly decreased in sepsis, such as IGLV1-5, IGLV4-69, IGLV7-43, and IGHV3-35. However, the immunoglobulins were removed from the list of the sepsis-related proteins because of their poor specificity.

Endothelial activation and vascular injury are one of the hallmarks of sepsis pathogenesis and were well reflected in the EV proteomic data. Various pathways related to endothelial activation leading to immune responses, such as neutrophil extracellular trap formation, actin cytoskeleton regulation, EC matrix-receptor interaction, and leukocyte trans-endothelial migration, were the most significant terms related to endothelial cell activation. DEFA1, DEFA3, S100A9, S100A8, CDC42, and TNN were differentially expressed in the septic EV cargos with strong AUROC and higher Z score when they were tested for sepsis diagnosis. Antimicrobial peptides, α-defensins, are constitutively expressed in neutrophils. Study found that the increased expression of DEFA1 (α-defensins-1) and DEFA3 (α-defensins-3) induces endothelial barrier dysfunction leading to more severe organ damage and mortality in mouse sepsis [32]. S100A8 and S100A9 proteins are involved in promoting endothelial cell activation through PI3K/Akt/mTOR pathway in human umbilical vein endothelial cells and are known as important regulators in cardiovascular inflammation [33]. Finally, we detected SERPINA1 and CDC42 with strong AUROC, which have been reported as potential sepsis biomarkers based on mRNA profiles [34]. As a regulatory factor in actin cytoskeleton and leukocyte trans-endothelial migration pathways, CDC42 contributes to vascular leakage and organ dysfunction in sepsis [35].

KEGG analysis of the M5 module revealed the upregulation of several metabolic pathways. The key words for those pathways within the 522 differential proteins are Proteasome, Carbon metabolism, Metabolic pathway, and Pyruvate metabolism. The discriminative EV proteins related metabolism in septic patients are MAPK4 (AUC=0.97), SLC16A3 (AUC=0.97), CP (AUC=0.96), and various histone proteins including H4C1 (AUC=0.99), H3C1 (AUC=0.95), and H2AX (AUC=0.95) in the upregulated candidates, while APOL1 (AUC=0.99) were detected in the downregulated proteins with strong AUROC and higher Z score. MAPK4 (Mitogen-activated protein kinase 4) is known as a noncanonical activator of AKT/mTOR signaling, which is related to metabolism [36]. SLC16A3 (Monocarboxylate transporter 4) is essential for the transport of lactate and pyruvate [37] and its role in sepsis is not well studied yet. CP (Ceruloplasmin) is reported to show a significant correlation with sepsis-related factors such as CRP and plays a role in liver dysfunction [38]. Various histone proteins, such as Histone H4 (H4C1), Histone H3.1 (H3C1), and Histone H2AX (H2AX) were significantly increased in septic EV proteome, except Histone H1.3 (H1-3), which decreased. The elevated extracellular histone levels are associated with the severity of sepsis and organ failure [39,40] especially H2B, H3, and H4. APOL1 is an important protein in lipid metabolism, facilitating lipid transfer and protecting against lipid oxidation. The downregulation of APOL1 is reported in septic patients showing reduced lipid metabolism as a significant change in sepsis [41].

In summary, using Tandem Mass Tag (TMT)-based quantitative proteomics, we profiled plasma EV proteins and performed a comprehensive bioinformatic analysis on these cargo proteins in septic patients and healthy controls. Our study identified many differentially expressed proteins in septic EVs, many of which were enriched in the functional pathways linked to sepsis pathogenesis, such as complement and coagulation, innate immunity, endothelial activation, and metabolic pathways, and possessed a strong predictive ability in sepsis diagnosis. These findings suggest the importance of EV cargo proteins in the host responses in sepsis and lay the foundation for future mechanistic study of EV proteins in sepsis.

## Methods

### Human subjects and blood collection

This study is part of an ongoing prospective observational study in septic patients conducted according to the policies and procedures as approved by the Institutional Review Board of the University of Maryland (HP-00081592). Informed consent was obtained for each septic patient and health control subject enrolled in the study. According to Sepsis-3 guidelines [2], the clinical criterion for sepsis is defined as infection plus a Sequential Organ Failure Assessment (SOFA) score ≥ 2. Septic patients between the ages of 18 and 80 years old with a SOFA ≥ 4 and suspected or confirmed infection upon admission to an ICU were enrolled. Patients with active malignancy or ongoing chemotherapy, critical illness not secondary to infection, or cardiac arrest prior to arrival were excluded. Age and sex-matched healthy (< 2 systemic conditions without functional impairment) control patients were recruited from the surrounding campus. Blood was collected in ethylenediaminetetraacetic acid (EDTA)-coated tubes within 24 hours of ICU admission and immediately processed for plasma preparation. The plasma samples were stored at −80°C until further analysis.

### Human plasma EV isolation

**Figure 1** illustrates the overall study design and workflow, as detailed below. Frozen plasma samples were thawed on ice to proceed with ultracentrifugation for EV isolation. Six hundred µL of plasma samples were diluted with the same volume of cold Dulbecco’s Phosphate Buffered Saline (DPBS) and centrifuged at 12,000 x *g* for 30 min at 4 °C. The supernatant was further diluted with 4 mL of cold DPBS and applied to ultracentrifugation at 110,000 x *g* for 1 hour at 4 °C (MLS50 fix-angle rotor, Beckman Coulter, Sykesville, MD, USA). We performed pilot experiments to determine the impact of washing steps on EV pellets based on the total proteins and the known exosome proteins recovered in the pellets after each step. These pilot data suggest that two washing steps after the initial ultracentrifugation – 3 rounds of ultracentrifugation - were optimal as they offered the best exosome protein recovery with minimal total protein loss (**Supplemental Table S1**). After the final pellet washing, the DPBS supernatant was removed, and the EV pellets were resuspended with 100 µL of cold DPBS. A small portion (5%) of EV samples was used for protein quantification and nanotracking particle analysis. For the subsequent proteomics analysis, the reconstituted EV samples were mixed with the same volume of 8 M Urea in 100 mM Tris buffer (pH 8). To detect EV markers on the EV samples, the western blot was performed with 25 µg of each EV sample using rabbit anti-Alix antibody (#18269, Cell Signaling Technology, MA, USA) and rabbit anti-CD9 antibody (#ab92726, Abcam).

### Sample preparation for Mass Spectrometry analysis

#### EV protein extraction and digestion

EV protein preparation for mass spectrometry analysis was conducted as described previously with minor modifications [42–44]. To extract proteins from plasma EVs, sample lysis was performed using ultrasonication (Branson sonifier 250, ultrasonics, Danbury, CT, USA) in 5 M urea and 50 mM Tris(hydroxymethyl)aminomethane hydrochloride (pH 8.0, Tris–HCl) for 1 min, with the samples maintained on the ice during the procedure. The protein was quantified using a bicinchoninic acid (BCA) assay kit (Pierce, Rockford, IL, USA). For reduction and alkylation, the samples were added with 10 mM Tris (2-carboxyethyl) phosphine hydrochloride (TCEP) and 40 mM chloroacetamide (CAA) to the final concentration. The mixture was incubated for 1 hour at room temperature. Subsequently, the protein digestion was initiated using LysC (Lysyl endopeptidase mass spectrometry grade; Fujifilm Wako Pure Chemical Industries Co., Ltd., Osaka, Japan) at a ratio of 1:100 for 3 h at 37 °C. This was followed by additional digestion with trypsin (sequencing grade modified trypsin; Promega, Fitchburg, WI, USA) at a ratio of 1:50 at 37 °C overnight. Prior to trypsin digestion, the urea concentration was reduced from 8 M to 2 M by adding three volumes of 50 mM triethylammonium bicarbonate (TEAB). The acidification of the samples was achieved with a final concentration of 1% trifluoroacetic acid (TFA), followed by desalting using C18 Stage-Tips (3M EmporeTM; 3M, St. Paul, MN, USA). The eluted peptide solution was subsequently subjected to vacuum drying using a Savant SPD121P SpeedVac concentrator (Thermo Scientific, Waltham, MA, USA).

#### Preparation of TMT-labeled EV peptides

To conduct quantitative mass spectrometry analysis, we labeled the prepared peptides with 2 batches of 14-plex TMTpro, a set of isobaric labels used for quantifying proteins from multiple samples simultaneously, according to the manufacturer’s instructions (Thermo Fisher Scientific). Briefly, TMT labeling reactions were conducted in acetonitrile (ACN) and 100 mM TEAB at room temperature for 1 h. The remaining free TMT tags were quenched by adding 100 mM Tris-HCl buffer (pH 8.0; Thermo Scientific) and incubating for over 5 min at room temperature. TMT-labeled EV peptides from each batch were pooled and dried using SpeedVac. For the normalization of data from 2 batches of the TMT experiment, we included the master pool (MP) in the last channel of each batch. The MP was prepared by mixing equal volumes from all 25 EV samples and was divided into each batch after completing the TMT labeling. To minimize the batch effect, the batch allocation and the order of 25 EV samples were block-randomized, keeping diagnosis, sex, and age balanced using an in-house R-script.

#### Preparation of TMT-labeled plasma peptides

Plasma samples were prepared in the same way as described for the plasma EV samples with the exception of the ultracentrifugation steps, which was omitted. These plasma proteins were enzymatically digested and labeled the same ways as described for EV protein samples.

#### Pre-fractionation

The peptides from each TMT experimental batch were subjected to basic pH reversed-phase liquid chromatography (bRPLC) fractionation on an Agilent 1260 High-Performance Liquid Chromatography (HPLC) system (Agilent Technologies, Santa Clara, CA, USA). This process included reconstituting peptides in solvent A (10 mM TEAB in water, pH 8.5), loading them onto an Agilent 300 Extend-C18 column (5 μm, 4.6 mm x 250 mm, Agilent Technologies), and resolving them over 90 minutes using a gradient of solvent B (10 mM TEAB in 90% ACN and 10% water, pH 8.5) at a flow rate of 0.3 mL/min. This led to the collection of 96 fractions, which were subsequently concatenated into 24 fractions and dried using a SpeedVac.

### LC-MS/MS

Liquid chromatography with tandem mass spectrometry (LC-MS/MS) analysis were conducted as described previously with minor modifications [42,43]. For the analysis involving LC-MS/MS, an Orbitrap Fusion Lumos Tribrid Mass Spectrometer (Thermo Fisher Scientific) was employed, coupled with an Ultimate 3000 RSLCnano nanoflow liquid chromatography system (Thermo Fisher Scientific). The peptides from each fraction were reconstituted in 0.5% formic acid (FA) and loaded onto a trap column (Acclaim PepMap 100, LC C18, 5 μm, 100 μm × 2 cm, nanoViper) at a flow rate of 8 μL/min. Subsequently, these peptides were separated at a flow rate of 0.3 μL/min through a gradient of solvent B (0.1% FA in 95% ACN) on an analytical column (Easy-Spray PepMap RSLC C18, 2 μm, 75 μm × 50 cm) equipped with an EASY-Spray ion source operating at a voltage of around 2.4 kV. The total duration of the run was 120 minutes. For MS analysis, the data-dependent acquisition (DDA) mode was employed, encompassing a full scan range of mass-to-charge ratio (*m/z*) 300 to 1800 in the “Top Speed” mode, with each cycle lasting 3 seconds. Both precursor mass (MS1) and fragment mass (MS2) scans were conducted for the precursor and the fragmentation ions of the peptide, respectively. MS1 scans were executed at a resolution of 120,000 at an *m/z* 200, while MS2 scans were carried out by fragmenting precursor ions using the higher-energy collisional dissociation (HCD) method, set to 35% of collision energy, and detected at a mass resolution of 50,000 at an *m/z* of 200. Automatic gain control targets were established at one million ions for MS1 and 0.05 million ions for MS2. The maximum ion injection time was set at 50 ms for MS1 and 100 ms for MS2. The precursor isolation window was defined as an *m/z* 1.6 with an offset of *m/z* 0.4. Dynamic exclusion was implemented for 30 seconds, and singly charged ions were rejected. Internal calibration was executed using the lock mass option (*m/z* 445.12002) derived from ambient air [45,46].

### Mass Spectrometry data

Data analysis was performed as described in our previous study with minor modifications [43]. The MS/MS data acquired from LC-MS/MS analyses were subjected to a database search using MSfragger 3.4 algorithm embedded in the Thermo Proteome Discoverer software package (version 2.4.1.15, Thermo Scientific) against the UniProt human protein database, which included both Swiss-Prot and TrEMBL entries (released in January 2019) and common contaminants [47]. During MS2 preprocessing, the top ten peaks within each 100 *m/z* window were chosen for database search. The following parameters were applied for the database search: Trypsin was designated as the protease, allowing for a maximum of two missed cleavages. Fixed modifications included carbamidomethylation of cysteine (+57.02146 Da) and TMT pro tags (+304.20715 Da) on lysine and peptide N termini. Methionine oxidation (+15.99492 Da) was considered as a variable modification. A minimum peptide length of six amino acids was set. MS1 and MS2 tolerances were established at 10 ppm and 20 ppm, respectively. False discovery rate (FDR) filtering was applied at 1% for both peptides and proteins using the percolator node and protein FDR validator node. For protein quantification, the integration mode employed the most confident centroid option, with a reporter ion tolerance of 20 ppm. The MS order was set to MS2, and HCD was selected as the activation type. Peptide quantification used both unique and razor peptides, while protein groups were considered for peptide uniqueness. The co-isolation threshold was set at 50%. Reporter ion abundance calculations were based on signal-to-noise ratios, with missing intensity values replaced with the minimum value. An average reporter signal-to-noise threshold of 10 was applied. Corrections for isobaric tags and data normalization were disabled. Protein grouping followed a strict parsimony principle, grouping proteins sharing the same set or subset of identified peptides. Protein groups lacking unique peptides were filtered out. The final protein groups were generated by iterating through all spectra, selecting PSMs with the highest number of unambiguous and unique peptides, and summing the reporter ion abundances of PSMs for corresponding proteins [48].

### Luminex multiplex assays

Luminex panels were designed with the following analytes from R&D Systems; 14 plex including IL6 (Interleukin6), TNFα (Tumor necrosis factor alpha), IL1β (Interleukin1 beta), IL8 (Interleukin8), CXCL2 (Chemokine (C-X-C motif) ligand 2), S100B (S100 calcium-binding protein B), Enolase2, Syndecan1, E-selectin, VCAM1 (Vascular cell adhesion protein 1), ICAM1 (Intercellular Adhesion Molecule 1, also known as CD54), Tie2, Angiopoietin1, and Angiopoietin2: 6 plex including vWF-A2 (Von Willebrand factor A2 domain), d-dimer, thrombomudulin, P-selectin, ADAMTS13 (a disintegrin and metalloproteinase with a thrombospondin type 1 motif, member 13), and Coagulation factor III/Tissue factor: and 3 plex including SerpinC1 (Antithrombin III), SerpinE1 (PAI-1), and CXCL4/PF4 (Platelet factor 4). Plasma samples were thawed on ice and centrifuged at 12,000 x g for 10 min. The supernatants were applied to the antibody-conjugated magnetic bead and proceeded further following the manufacturer’s instructions. The fluorescent antibody reaction was measured on a Luminex 200 machine and the analysis was performed with the Luminex xPONENT software.

### Statistical and bioinformatics

#### Experimental design and statistical rationale

Experimental design and statistical analyses were performed as described previously with minor modifications [42,43]. The total number of plasma-derived EV samples used in this study was 25, composed of 15 septic patients and 10 healthy controls. These numbers were determined based on our power analysis using the pwr package in R [42]. Since our goal was to detect proteins with 1.5-fold differences between groups, the required minimum sample size was 7.934 when the significance level was 0.0003, power was 0.8, sigma was 0.2, and delta was 0.585 (=log_2_1.5). This sigma value of 0.2 was derived from our in-house TMT proteomics experiments. The significance level of 0.0003 was determined based on our in-house data. When we identified approximately thousands of proteins, a majority of the proteins with *P* value < 0.0003 showed q-value < 0.05. Based on this sample size analysis, we decided to use 10 or more samples per group. The statistical analysis of the mass spectrometry data was conducted using the Perseus software package (version 1.6.15.0) [49]. For normalization, the reporter ion intensity values were divided by the MP values of each protein, and the relative abundance values for each sample were subtracted by the median values of each sample after the log_2_ transformation. Subsequently, a z-score transformation was applied to the data. The *P* values between the comparison groups were calculated using the student’s two-sample t-test, because the protein abundances showed normal distribution, according to our normality test using the Shapiro–Wilk test in the dplyr package in R [50]. Proteins with q-values < 0.05 were considered differentially expressed. Given the context of handling multiple comparisons, the FDR was calculated by comparing data with and without permutations between groups. The q-values for the volcano plot were calculated by significance analysis of microarray (SAM) and a permutation-based FDR estimation with 0.1 of the S0 value [51].

#### Bioinformatic analysis

The proteins that displayed differential expression were employed in the Kyoto Encyclopedia of Genes and Genomes (KEGG) pathway and Gene Ontology (GO) analysis embedded in DAVID bioinformatics resources (version 6.8) [52]. The gene set enrichment was performed using a GO plot [53] to visualize the KEGG pathway results using the package in R. For interactome analysis, the STRING Protein-Protein Interaction (PPI) database version 11 was utilized [54]. Additionally, the weighted gene co-expression network analysis (WGCNA) was performed using the R software package [55]. Receiver operating characteristic (ROC) curve and Principal Component Analysis (PCA) data were generated using the MetaboAnalyst tool [56]. Additionally, for the cell-type database, we employed a database of Deeply Integrated human Single-Cell Omics data (DISCO), which provides information on marker proteins for 461 cell types derived from single-cell RNA-seq data [57]. The *P* values between the cell-type database and WGCNA modules were evaluated using the Fisher exact test [58]. Pearson correlation was calculated between the plasma and EV proteomes, and the results were visualized using histograms.

## Data availability

All MS data and search results have been deposited to the ProteomeXchange Consortium (https://www.proteomexchange.org) via the PRIDE partner repository with the dataset identifier PXD056258. Reviewers can access the dataset by using ‘’ as the ID and ‘s9p3QJWHn81m’ as the password.

## List of abbreviations

The abbreviations used are:

EVs: extracellular vesicles
HC: healthy control
TMT: tandem mass tag
WGCNA: Weighted gene coexpression network analysis
KEGG: Kyoto Encyclopedia of Genes and Genomes
GO: Gene Ontology
SOFA: sequential organ failure assessment
EDTA: Ethylenediaminetetraacetic acid
DPBS: Dulbecco’s Phosphate Buffered Saline
Tris-HCl: Tris(hydroxymethyl)aminomethane hydrochloride
BCA: bicinchoninic acid
TCEP: tris(2-carboxyethyl) phosphine hydrochloride
CAA: chloroacetamide
TEAB: triethylammonium bicarbonate
TFA: trifluoroacetic acid
ACN: acetonitrile
MP: master pool
bRPLC: basic pH reversed-phase liquid chromatography
HPLC: High-Performance Liquid Chromatography
LC-MS/MS: Liquid chromatography with tandem mass spectrometry
FA: formic acid
DDA: data-dependent acquisition
m/z: mass-to-charge ratio
MS1: precursor mass
MS2: fragment mass
HCD: higher-energy collisional dissociation
FDR: False discovery rate
SAM: Significance Analysis of Microarrays
PPI: Protein-Protein Interaction
ROC: Receiver operating characteristic
PCA: Principal Component Analysis
DISCO: Deeply Integrated human Single-Cell Omics
NTA: nanoparticle tracking analysis
GO-CC: Gene Ontology Cellular Components
EC: endothelial cell
MM: Module Membership
PS: Protein Significance
BUN: blood urea nitrogen
Cr: creatinine
PLT: platelet count
INR: international normalized ratio
PTT: partial thromboplastin
PT: prothrombin time
WBC: White blood cell count
Hg: hemoglobin
Hct: hematocrit
K: potassium
Na: sodium
PRM: Parallel Reaction Monitoring.

## Authors’ contributions

CP, TR, CN, and WC conceptualized and designed the project. RM-H and BW recruited patients and collected plasma samples. CP and TR performed EV isolation and characterization and mass spectrometry analysis. TR, KK, and JK analyzed the mass spectrometry data and conducted bioinformatic analysis. CP, TR, CN, and WC wrote and edited the manuscript. CN and WC supervised the project.

## Acknowledgments

We acknowledge an NIH shared instrumentation grant (S10OD021844). Figure 1 was created with BioRender.com.

## Declarations

This research study was approved by the Institutional Review Board (HP 00081592), Maryland University Baltimore (UMB). This study abided by the Declaration of Helsinki principles.

## Consent for publication

All authors consent to the publication of this manuscript.

## References

1. Coopersmith, C.M., et al. Surviving sepsis campaign: Research priorities for sepsis and septic shock. Intensive Care Med 44, 1400–1426 (2018).

2. Singer, M., et al. The third international consensus definitions for sepsis and septic shock (sepsis-3). JAMA 315, 801–810 (2016).

3. Fleischmann-Struzek, C., et al. Incidence and mortality of hospital- and icu-treated sepsis: Results from an updated and expanded systematic review and meta-analysis. Intensive Care Med 46, 1552–1562 (2020).

4. Schlapbach, L.J., et al. International consensus criteria for pediatric sepsis and septic shock. JAMA 331, 665–674 (2024).

5. Yanez-Mo, M., et al. Biological properties of extracellular vesicles and their physiological functions. J Extracell Vesicles 4, 27066 (2015).

6. Ciferri, M.C., Quarto, R. & Tasso, R. Extracellular vesicles as biomarkers and therapeutic tools: From pre-clinical to clinical applications. Biology (Basel) 10(2021).

7. Wiklander, O.P.B., Brennan, M.A., Lotvall, J., Breakefield, X.O. & El Andaloussi, S. Advances in therapeutic applications of extracellular vesicles. Sci Transl Med 11(2019).

8. Sanwlani, R. & Gangoda, L. Role of extracellular vesicles in cell death and inflammation. Cells 10(2021).

9. Raeven, P., Zipperle, J. & Drechsler, S. Extracellular vesicles as markers and mediators in sepsis. Theranostics 8, 3348–3365 (2018).

10. Tian, C., et al. Extracellular vesicles participate in the pathogenesis of sepsis. Front Cell Infect Microbiol 12, 1018692 (2022).

11. Li, P., et al. Circulating extracellular vesicles are associated with the clinical outcomes of sepsis. Front Immunol 14, 1150564 (2023).

12. Plaschke, K., et al. Extracellular vesicles as possible plasma markers and mediators in patients with sepsis-associated delirium-a pilot study. Int J Mol Sci 24(2023).

13. Thompson, A., et al. Tandem mass tags: A novel quantification strategy for comparative analysis of complex protein mixtures by ms/ms. Anal Chem 75, 1895–1904 (2003).

14. Kim, J., Lee, H., Park, K. & Shin, S. Rapid and efficient isolation of exosomes by clustering and scattering. J Clin Med 9(2020).

15. Jang, Y., et al. Mass spectrometry-based proteomics analysis of human substantia nigra from parkinson’s disease patients identifies multiple pathways potentially involved in the disease. Mol Cell Proteomics 22, 100452 (2023).

16. Williams, B., Zou, L., Pittet, J.F. & Chao, W. Sepsis-induced coagulopathy: A comprehensive narrative review of pathophysiology, clinical presentation, diagnosis, and management strategies. Anesth Analg 138, 696–711 (2024).

17. Arora, J., Mendelson, A.A. & Fox-Robichaud, A. Sepsis: Network pathophysiology and implications for early diagnosis. Am J Physiol Regul Integr Comp Physiol 324, R613–R624 (2023).

18. Futosi, K., Fodor, S. & Mocsai, A. Neutrophil cell surface receptors and their intracellular signal transduction pathways. Int Immunopharmacol 17, 638–650 (2013).

19. Yuki, K. & Hou, L. Role of beta2 integrins in neutrophils and sepsis. Infect Immun 88(2020).

20. Kremer Hovinga, J.A., et al. Adamts-13, von willebrand factor and related parameters in severe sepsis and septic shock. J Thromb Haemost 5, 2284–2290 (2007).

21. van Mourik, J.A., et al. Von willebrand factor propeptide in vascular disorders: A tool to distinguish between acute and chronic endothelial cell perturbation. Blood 94, 179–185 (1999).

22. Sarani, N., et al. Clinical utility of recently food and drug administration-approved intellisep test (sepsis biomarker) for early diagnosis of sepsis: Comparison with other biomarkers. J Clin Med 13(2024).

23. Levi, M. & van der Poll, T. Coagulation and sepsis. Thromb Res 149, 38–44 (2017).

24. Botia, M., et al. Gaining knowledge about biomarkers of the immune system and inflammation in the saliva of pigs: The case of myeloperoxidase, s100a12, and itih4. Res Vet Sci 164, 104997 (2023).

25. Donaldson, J.G. & Jackson, C.L. Arf family g proteins and their regulators: Roles in membrane transport, development and disease. Nat Rev Mol Cell Biol 12, 362–375 (2011).

26. Zhou, Q., Hao, L., Huang, W. & Cai, Z. The golgi-associated plant pathogenesis-related protein gapr-1 enhances type i interferon signaling pathway in response to toll-like receptor 4. Inflammation 39, 706–717 (2016).

27. Olesch, C., Ringel, C., Brune, B. & Weigert, A. Beyond immune cell migration: The emerging role of the sphingosine-1-phosphate receptor s1pr4 as a modulator of innate immune cell activation. Mediators Inflamm 2017, 6059203 (2017).

28. Cummings, C.J., et al. Expression and function of the chemokine receptors cxcr1 and cxcr2 in sepsis. J Immunol 162, 2341–2346 (1999).

29. Randhawa, A.K., Ziltener, H.J. & Stokes, R.W. Cd43 controls the intracellular growth of mycobacterium tuberculosis through the induction of tnf-alpha-mediated apoptosis. Cell Microbiol 10, 2105–2117 (2008).

30. Mizuno, S. & Nakamura, T. Improvement of sepsis by hepatocyte growth factor, an anti-inflammatory regulator: Emerging insights and therapeutic potential. Gastroenterol Res Pract 2012, 909350 (2012).

31. Zhou, Y., et al. Usefulness of the heparin-binding protein level to diagnose sepsis and septic shock according to sepsis-3 compared with procalcitonin and c reactive protein: A prospective cohort study in china. BMJ Open 9, e026527 (2019).

32. Chen, Q., et al. Increased gene copy number of defa1/defa3 worsens sepsis by inducing endothelial pyroptosis. Proc Natl Acad Sci U S A 116, 3161–3170 (2019).

33. Sun, Y., et al. S100a8/a9 proteins: Critical regulators of inflammation in cardiovascular diseases. Front Cardiovasc Med 11, 1394137 (2024).

34. Yang, Y.X. & Li, L. Identification of potential biomarkers of sepsis using bioinformatics analysis. Exp Ther Med 13, 1689–1696 (2017).

35. Chen, M., et al. The circulating cdc42 expression in sepsis: Relation to disease susceptibility, inflammation, multiple organ dysfunctions and mortality risk. Ann Clin Lab Sci 54, 525–532 (2024).

36. Wang, W., et al. Mapk4 overexpression promotes tumor progression via noncanonical activation of akt/mtor signaling. J Clin Invest 129, 1015–1029 (2019).

37. Felmlee, M.A., Jones, R.S., Rodriguez-Cruz, V., Follman, K.E. & Morris, M.E. Monocarboxylate transporters (slc16): Function, regulation, and role in health and disease. Pharmacol Rev 72, 466–485 (2020).

38. Chiarla, C., Giovannini, I. & Siegel, J.H. Patterns of correlation of plasma ceruloplasmin in sepsis. J Surg Res 144, 107–110 (2008).

39. Garcia-Gimenez, J.L., et al. Validation of circulating histone detection by mass spectrometry for early diagnosis, prognosis, and management of critically ill septic patients. J Transl Med 21, 344 (2023).

40. Xu, J., et al. Extracellular histones are major mediators of death in sepsis. Nat Med 15, 1318–1321 (2009).

41. Sharma, N.K., et al. Lipid metabolism impairment in patients with sepsis secondary to hospital acquired pneumonia, a proteomic analysis. Clin Proteomics 16, 29 (2019).

42. Jang, Y., et al. Mass spectrometry-based proteomics analysis of human globus pallidus from progressive supranuclear palsy patients discovers multiple disease pathways. Clin Transl Med 12, e1076 (2022).

43. Ryu, T., Kim, S.Y., Thuraisamy, T., Jang, Y. & Na, C.H. Development of an in situ cell-type specific proteome analysis method using antibody-mediated biotinylation. bioRxiv (2023).

44. Soldan, A., et al. Nptx2 in cerebrospinal fluid predicts the progression from normal cognition to mild cognitive impairment. Ann Neurol 94, 620–631 (2023).

45. Khan, S.Y., et al. Examining the effects of cigarette smoke on mouse lens through a multi omic approach. Sci Rep 11, 18801 (2021).

46. Khan, M.R., et al. Enhanced mtorc1 signaling and protein synthesis in pathologic alpha-synuclein cellular and animal models of parkinson’s disease. Sci Transl Med 15, eadd0499 (2023).

47. Kong, A.T., Leprevost, F.V., Avtonomov, D.M., Mellacheruvu, D. & Nesvizhskii, A.I. Msfragger: Ultrafast and comprehensive peptide identification in mass spectrometry-based proteomics. Nat Methods 14, 513–520 (2017).

48. Ramachandran, K.V., et al. Activity-dependent degradation of the nascentome by the neuronal membrane proteasome. Mol Cell 71, 169–177 e166 (2018).

49. Tyanova, S., et al. The perseus computational platform for comprehensive analysis of (prote)omics data. Nat Methods 13, 731–740 (2016).

50. Shapiro, S.S. & Wilk, M.B. An analysis of variance test for normality (complete samples). Biometrika 52, 591–611 (1965).

51. Tusher, V.G., Tibshirani, R. & Chu, G. Significance analysis of microarrays applied to the ionizing radiation response. Proc Natl Acad Sci U S A 98, 5116–5121 (2001).

52. Huang da, W., Sherman, B.T. & Lempicki, R.A. Systematic and integrative analysis of large gene lists using david bioinformatics resources. Nat Protoc 4, 44–57 (2009).

53. Walter, W., Sanchez-Cabo, F. & Ricote, M. Goplot: An r package for visually combining expression data with functional analysis. Bioinformatics 31, 2912–2914 (2015).

54. Szklarczyk, D., et al. String v11: Protein-protein association networks with increased coverage, supporting functional discovery in genome-wide experimental datasets. Nucleic Acids Res 47, D607–D613 (2019).

55. Langfelder, P. & Horvath, S. Wgcna: An r package for weighted correlation network analysis. BMC Bioinformatics 9, 559 (2008).

56. Pang, Z., et al. Using metaboanalyst 5.0 for lc-hrms spectra processing, multi-omics integration and covariate adjustment of global metabolomics data. Nat Protoc 17, 1735–1761 (2022).

57. Li, M., et al. Disco: A database of deeply integrated human single-cell omics data. Nucleic Acids Res 50, D596–D602 (2022).

58. Johnson, E.C.B., et al. Large-scale deep multi-layer analysis of alzheimer’s disease brain reveals strong proteomic disease-related changes not observed at the rna level. Nat Neurosci 25, 213–225 (2022).

59. Wang, L., Bastarache, J.A. & Ware, L.B. The coagulation cascade in sepsis. Curr Pharm Des 14, 1860–1869 (2008).

60. Garcia-Obregon, S., et al. Identification of a panel of serum protein markers in early stage of sepsis and its validation in a cohort of patients. J Microbiol Immunol Infect 51, 465–472 (2018).

61. Tang, J., et al. Propofol lowers serum pf4 level and partially corrects hypercoagulopathy in endotoxemic rats. Biochim Biophys Acta 1804, 1895–1901 (2010).

62. Wang, M., Zhong, D., Dong, P. & Song, Y. Blocking cxcr1/2 contributes to amelioration of lipopolysaccharide-induced sepsis by downregulating substance p. J Cell Biochem 120, 2007–2014 (2019).

63. Yuan, W., et al. Clinical significance and prognosis of serum tenascin-c in patients with sepsis. BMC Anesthesiol 18, 170 (2018).

