## Supplemental Figures for "Proteomic profiling of plasma extracellular vesicles identifies signatures of innate immunity, coagulation, and endothelial activation in septic patients"

^1^Translational Research Program, Department of Anesthesiology, University of Maryland School of Medicine, Baltimore, MD, USA; ^2^Center for Shock, Trauma and Anesthesiology Research, University of Maryland School of Medicine, Baltimore, MD, USA; ^3^Department of Neurology, Johns Hopkins University School of Medicine, Baltimore, MD, USA; ^4^Neuroregeneration and Stem Cell Programs, Institute for Cell Engineering, Johns Hopkins University School of Medicine, Baltimore, MD, USA.

^#^These authors contributed equally.

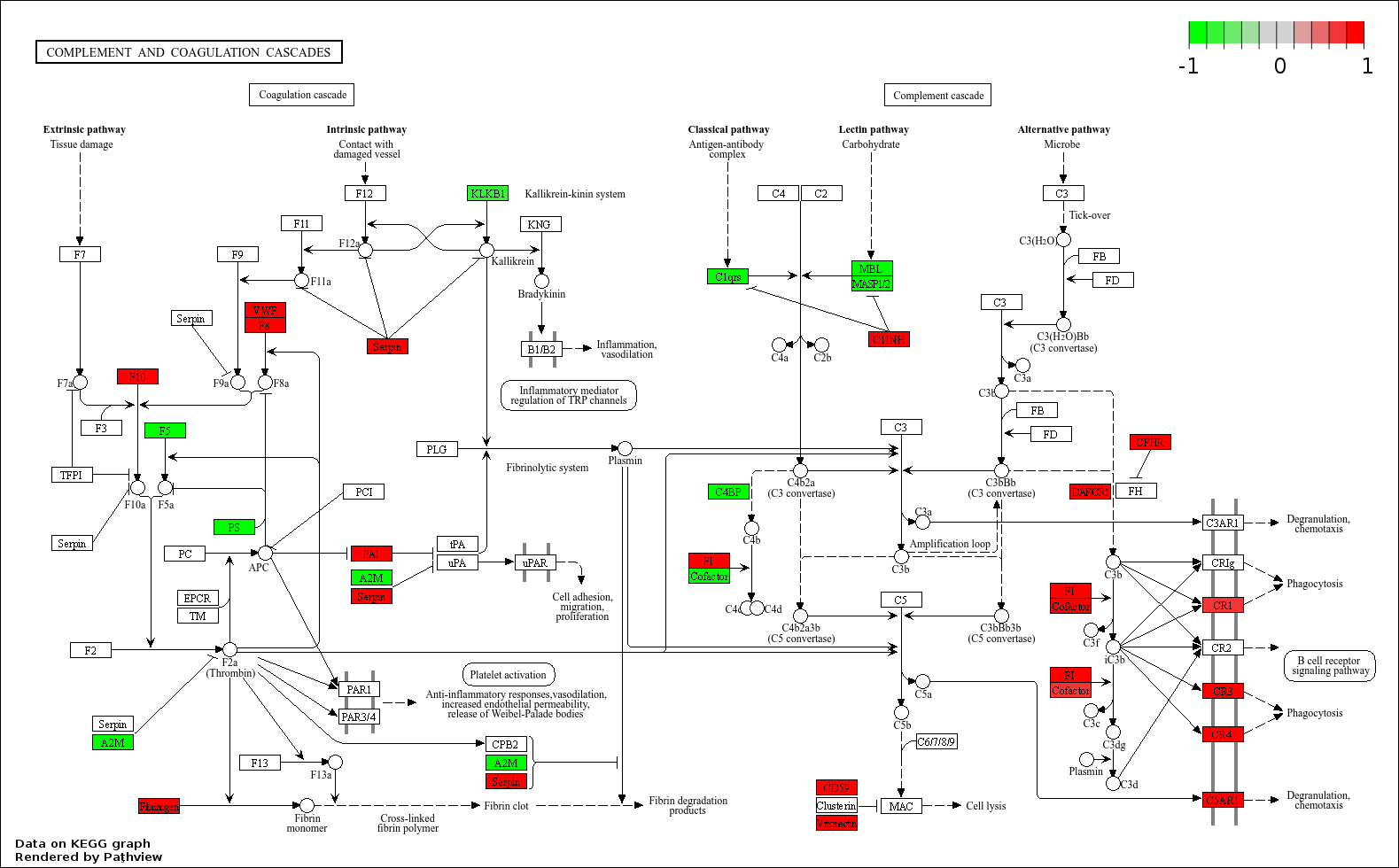

**Figure S1. An illustration of the complement and coagulation cascades (homosapiens) from KEGG map.**

This map displays the pathway of 30 out of 138 proteins in the complement and coagulation systems, which were the most enriched according to the KEGG results. Proteins with significant down- and up-regulation in the septic EV cargo are shown in green and red, respectively. This illustration shows the expression of genes mapping to the complement and coagulation cascades in the KEGG map and is displayed using the Pathview R package.

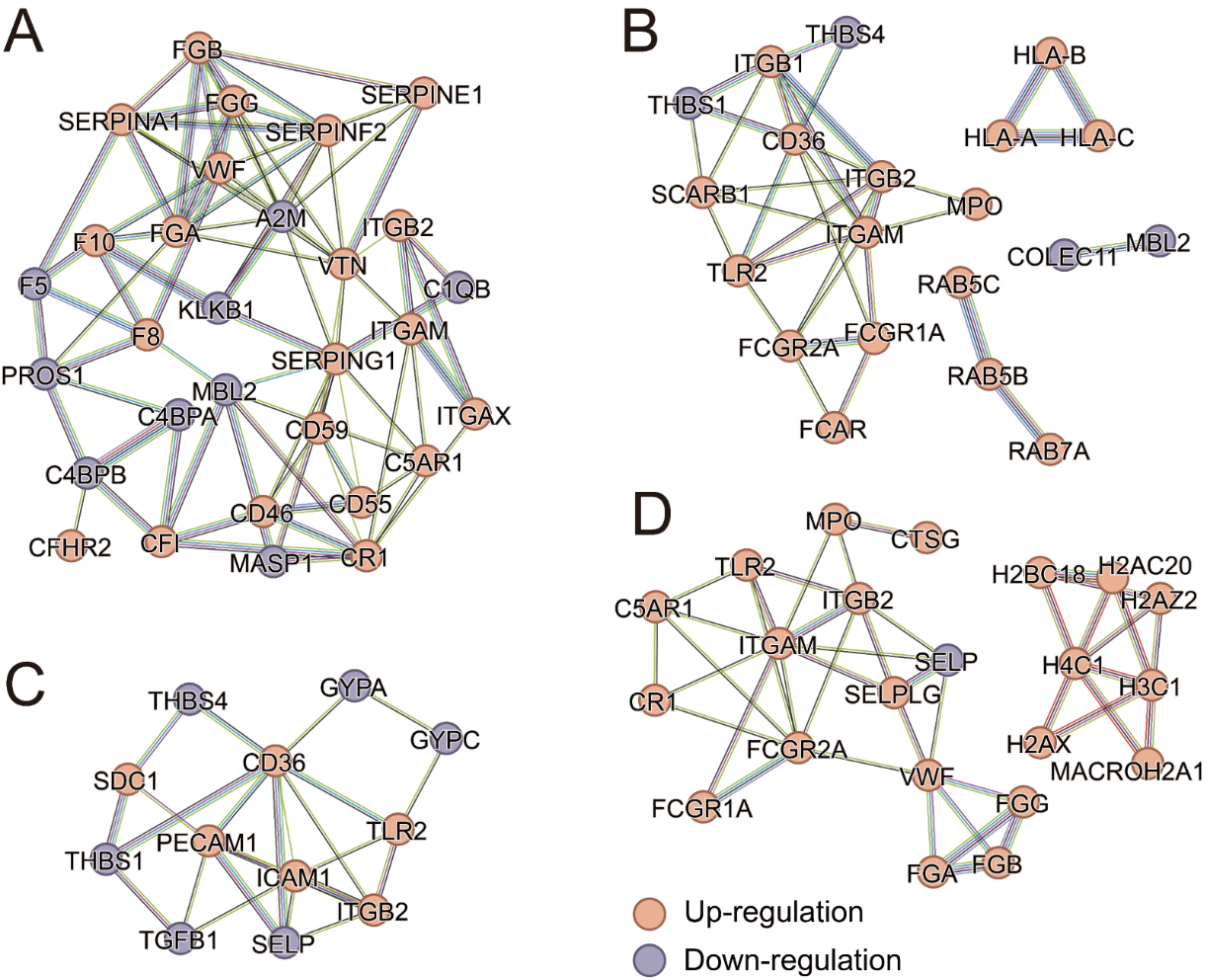

**Figure S2. STRING PPI analysis of proteins from the top 4 most enriched KEGG pathways among the differentially expressed EV proteins in septic patients.**

(A) Protein-protein interaction network analysis of proteins enriched in Complement and Coagulation Cascades using STRING. This STRING PPI analysis result contains 30 nodes with 81 edges. Experimental and database evidence was used for the active interaction source with the high confidence score of 0.7 as the minimum required interaction score (average node degree: 5.4, average local clustering coefficient: 0.449, and PPI enrichment *P* < 1.0e^-16^).  (B) Protein-protein interaction network analysis of proteins enriched in Phagosome using STRING. This STRING PPI analysis result contains 22 nodes with 34 edges. Experimental and database evidence was used for the active interaction source with the high confidence score of 0.7 as the minimum required interaction score (average node degree: 3.09, average local clustering coefficient: 0.676, and PPI enrichment *P* < 1.0e^-16^).  (C) Protein-protein interaction network analysis of proteins enriched in Malaria using STRING. This STRING PPI analysis result contains 13 nodes with 24 edges. Experimental and database evidence was used for the active interaction source with the high confidence score of 0.7 as the minimum required interaction score (average node degree: 3.69, average local clustering coefficient: 0.307, and PPI enrichment *P* < 1.0e^-16^).  (D) Protein-protein interaction network analysis of proteins enriched in Neutrophil extracellular trap formation using STRING. This STRING PPI analysis result contains 24 nodes with 43 edges. Experimental and database evidence was used for the active interaction source with the high confidence score of 0.7 as the minimum required interaction score (average node degree: 3.58, average local clustering coefficient: 0.649, and PPI enrichment *P* < 1.0e^-16^). The disconnected nodes have been removed from the network. Edges with yellow, purple, light blue, black, green, red, and blue colors represent protein-protein interaction evidence based on text-mining, experimental, database, co-expression, neighborhood, gene fusion, and co-occurrence evidence, respectively.

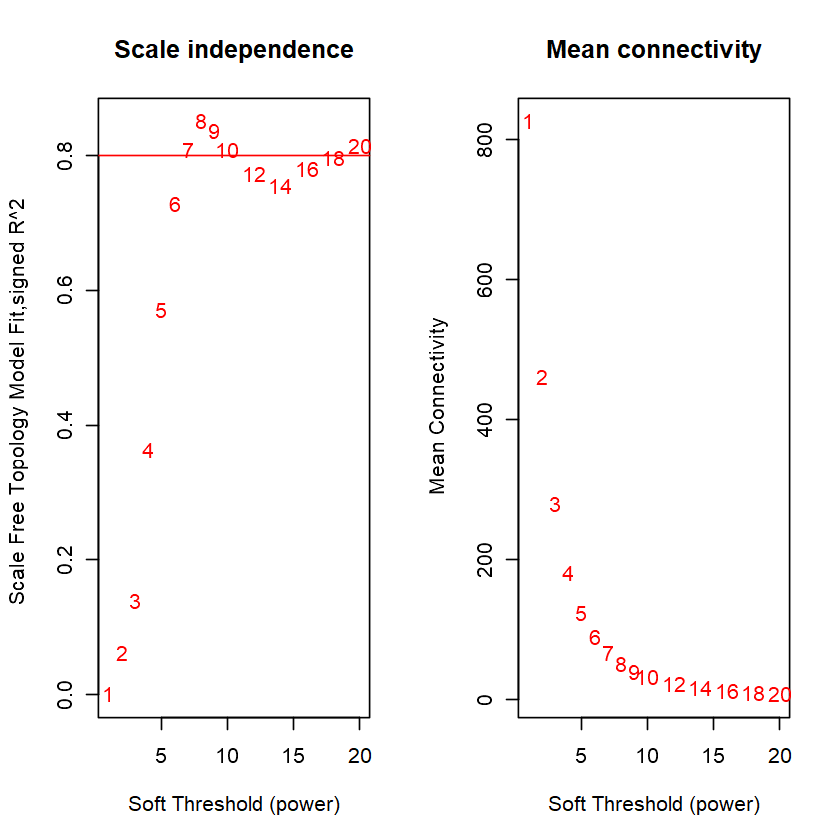

**Figure S3. Scale independence and mean connectivity in WGCNA.**

Determining soft-thresholding power in WGCNA: the scale-free fit index and the mean connectivity for various soft-thresholding powers.

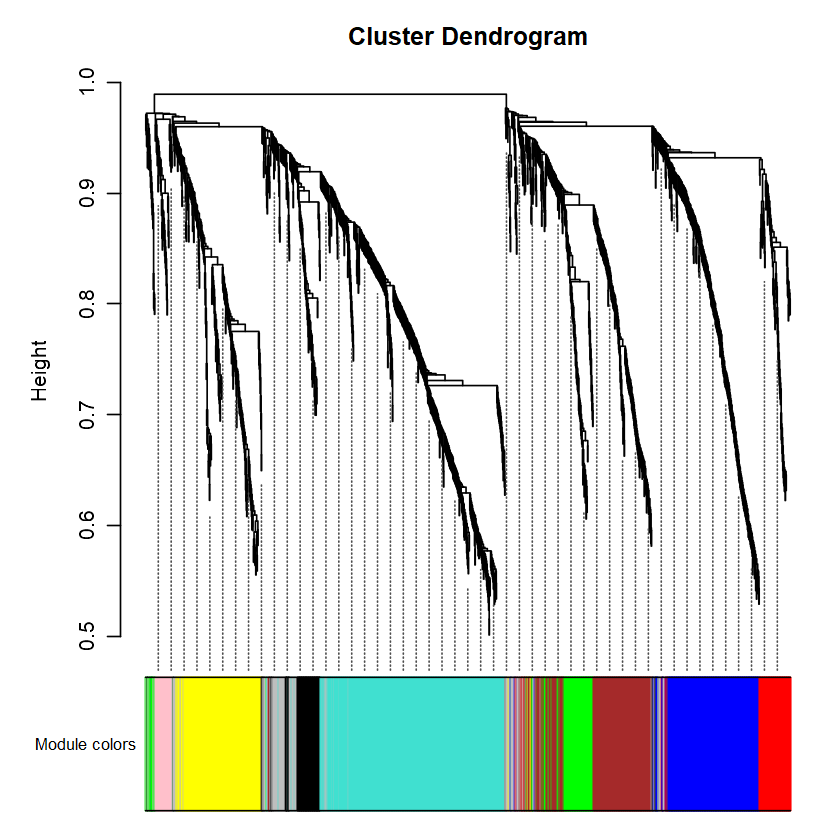

**Figure S4. Cluster dendrogram in WGCNA.**

A cluster dendrogram was created by WGCNA of the EV proteome data from septic patients and HC individuals to identify protein modules that show co-expression patterns.

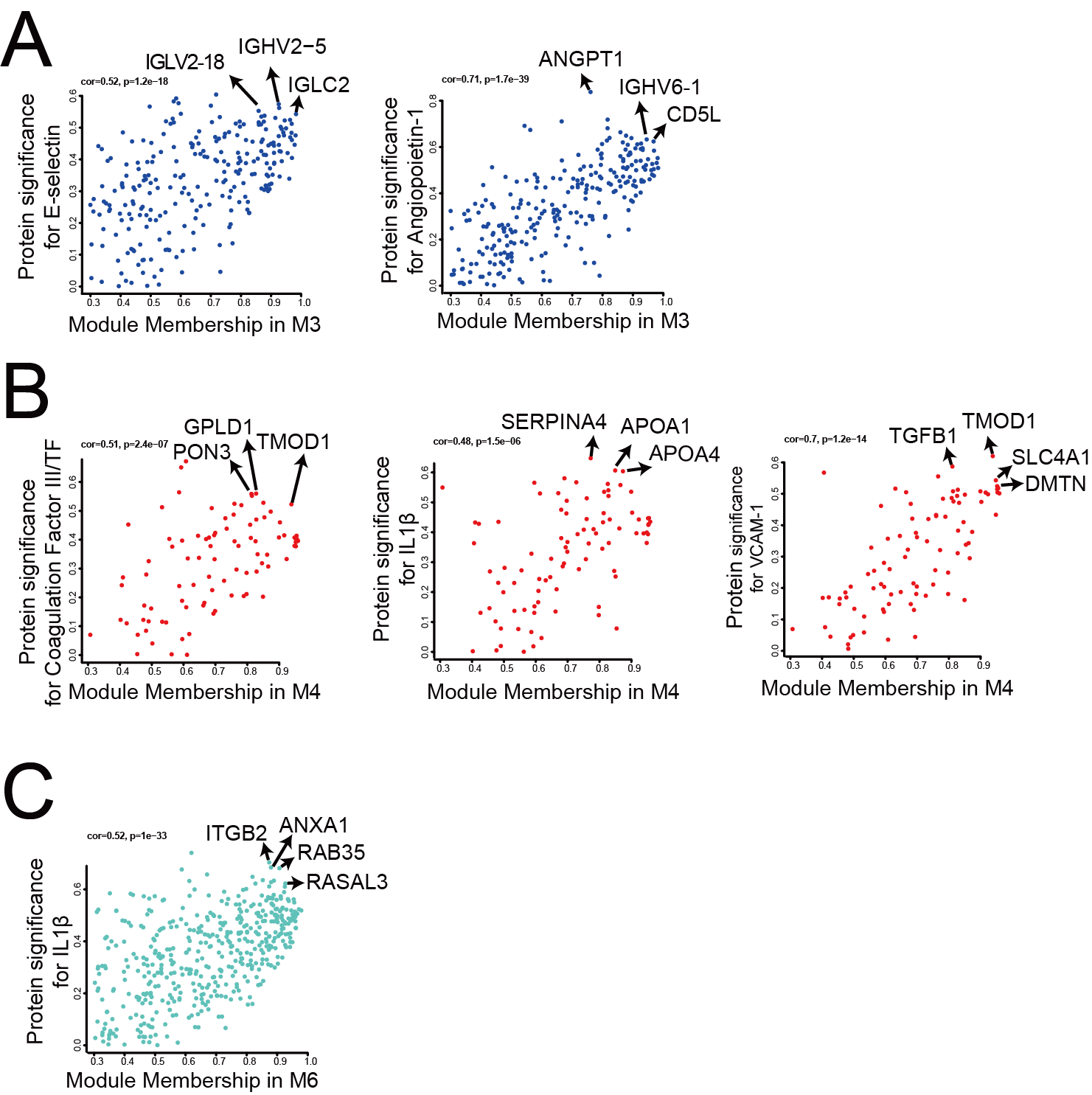

**Figure S5. Module membership (MM) and protein significance (PS) analysis.**

(A, B, C) MM-PS plots showing the relationship between MM and PS for proteins that were not among the top 5 traits but still exhibited significance at *P* < 0.01 in the M3 (A), M4 (B), and M6 (C) modules from Figure 5.

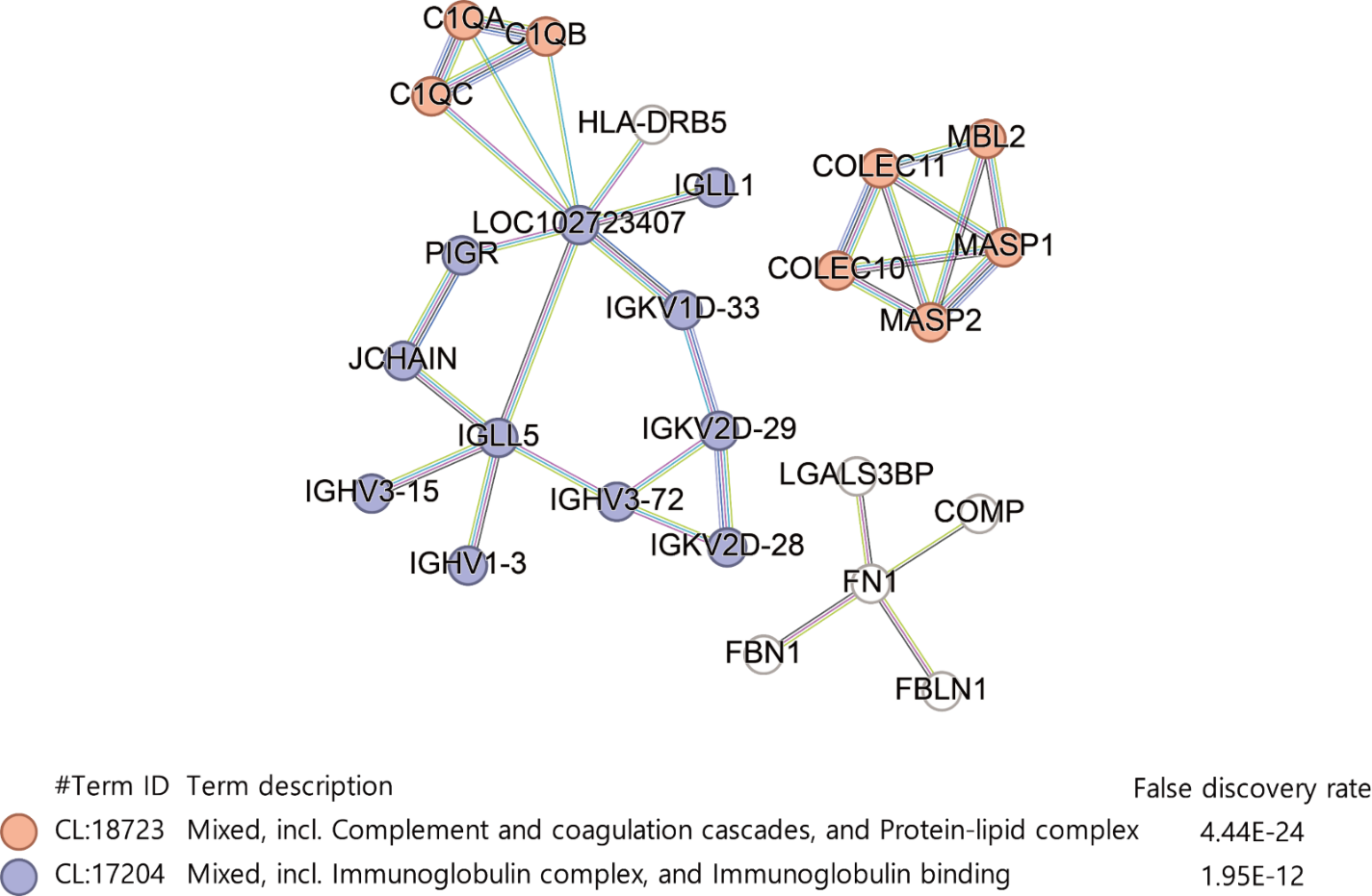

**Figure S6. STRING PPI analysis of the proteins in the M3 module.**

This STRING PPI analysis result contains 25 nodes with 33 edges. The minimum required confidence score was set to 0.9 (average node degree: 0.622, average local clustering coefficient: 0.245, and PPI enrichment P < 4.71e-12). The networks formed with four or fewer nodes were removed. Edges with green, blue, purple, yellow, light blue, and black colors represent the protein-protein interaction evidence based on neighborhood, co-occurrence, experimental, text mining, database, and co-expression, respectively.

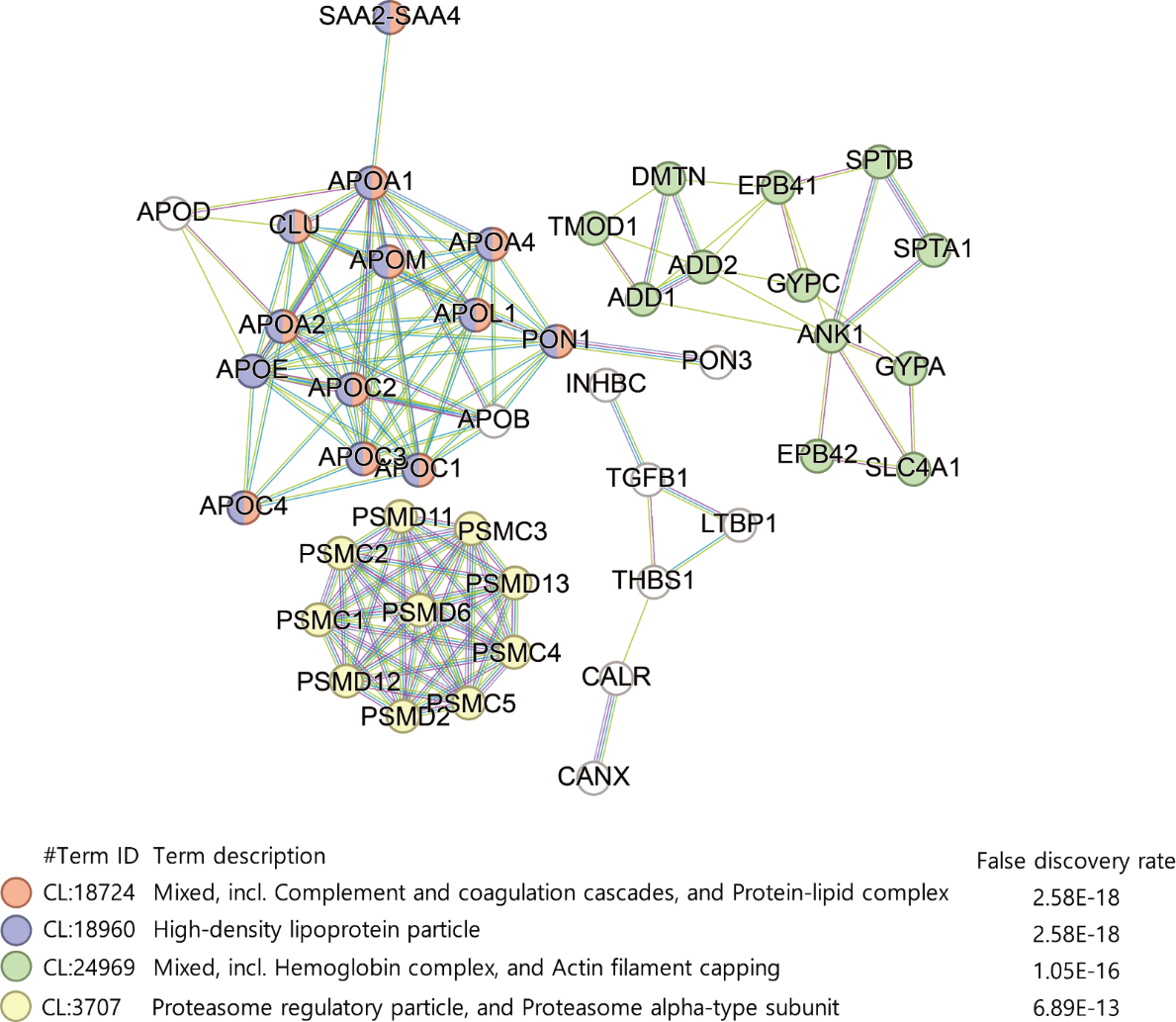

**Figure S7. STRING PPI analysis of the proteins in the M4 module.**

This STRING PPI analysis result contains 82 nodes with 147 edges. The minimum required confidence score was set to 0.9 (average node degree: 3.36, average local clustering coefficient: 0.483, and PPI enrichment P < 1.0e-16). The networks formed with four or fewer nodes were removed. The networks formed with four or fewer nodes were removed. Protein-protein interaction analysis was conducted using STRING based on the parameters of text mining (yellow edges), experimental evidence (purple edges), and database information (blue edges).

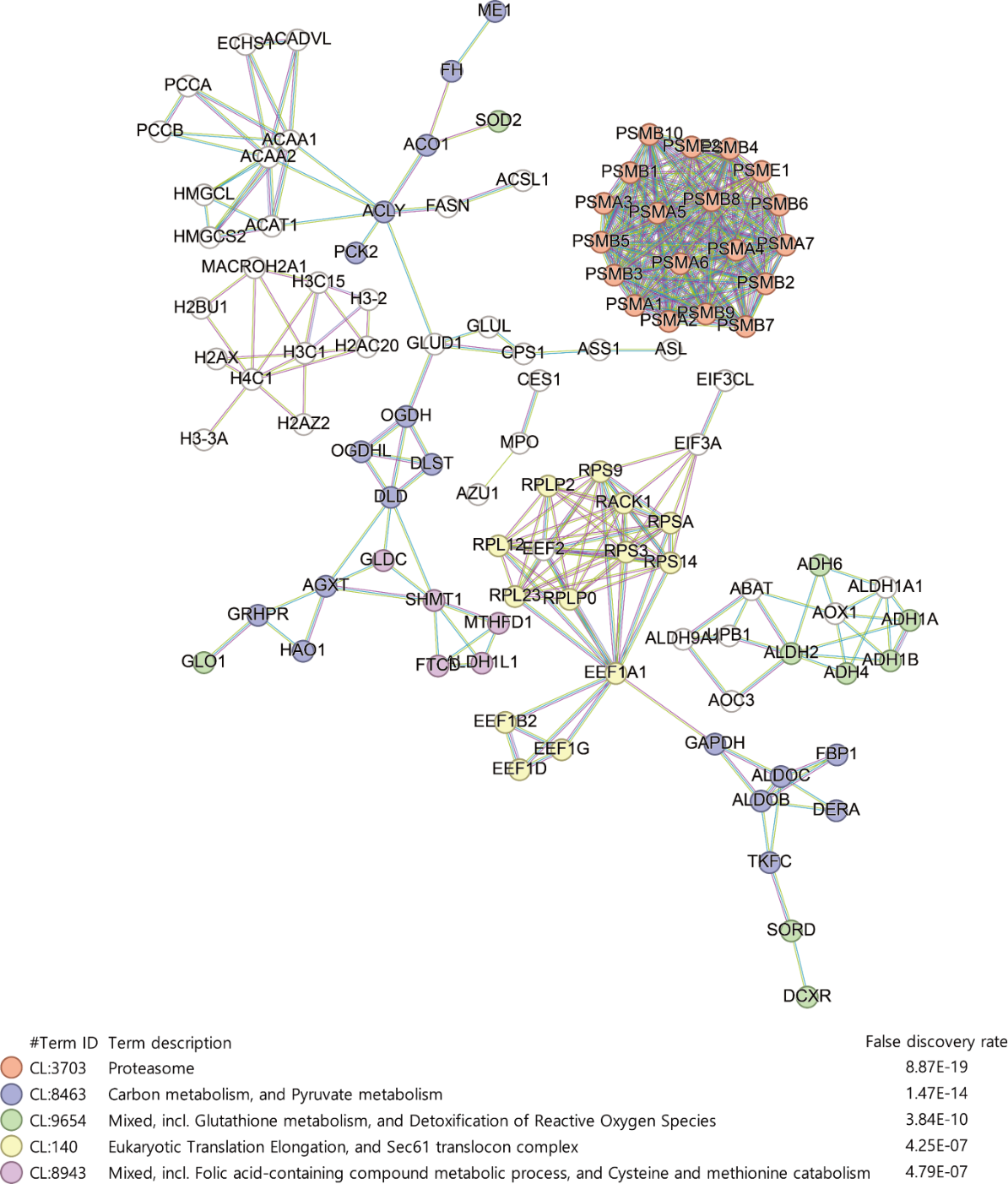

**Figure S8. STRING PPI analysis of the proteins in the M5 module.**

This STRING PPI analysis result contains 188 nodes with 347 edges. The minimum required confidence score was set to 0.9 (average node degree: 3.46, average local clustering coefficient: 0.445, and PPI enrichment P < 1.0e-16). The networks formed with four or fewer nodes were removed. Protein-protein interaction analysis was conducted using STRING based on the parameters of text mining (yellow edges), experimental evidence (purple edges), and database information (blue edges).

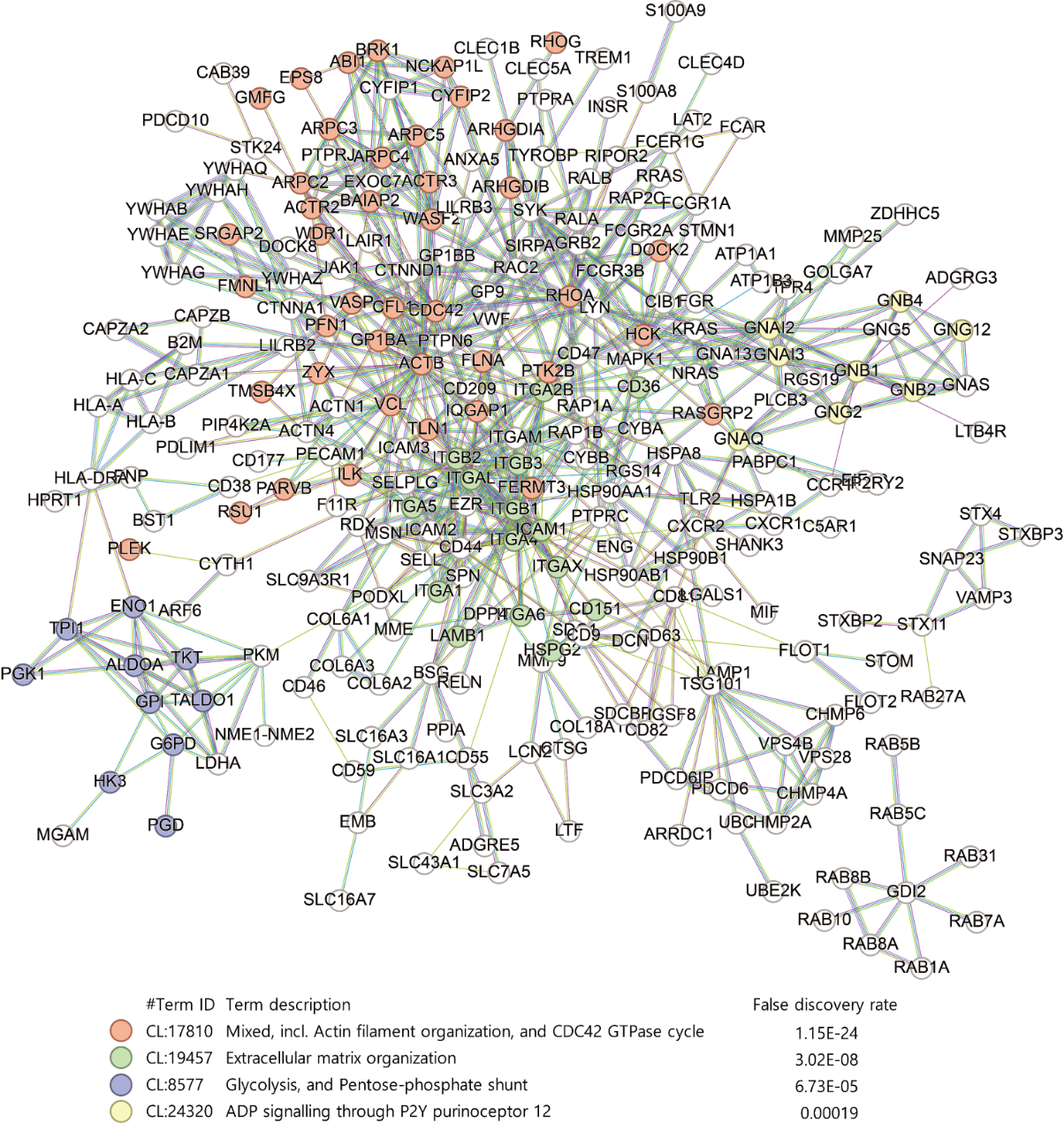

**Figure S9. STRING PPI analysis of the proteins in the M6 module.**

This STRING PPI analysis result contains 428 nodes with 741 edges. The minimum required confidence score was set to 0.9 (average node degree: 3.3, average local clustering coefficient: 0.414, and PPI enrichment P < 1.0e-16). The networks formed with four or fewer nodes were removed. Protein-protein interaction analysis was conducted using STRING based on the parameters of text mining (yellow edges), experimental evidence (purple edges), and database information (blue edges).

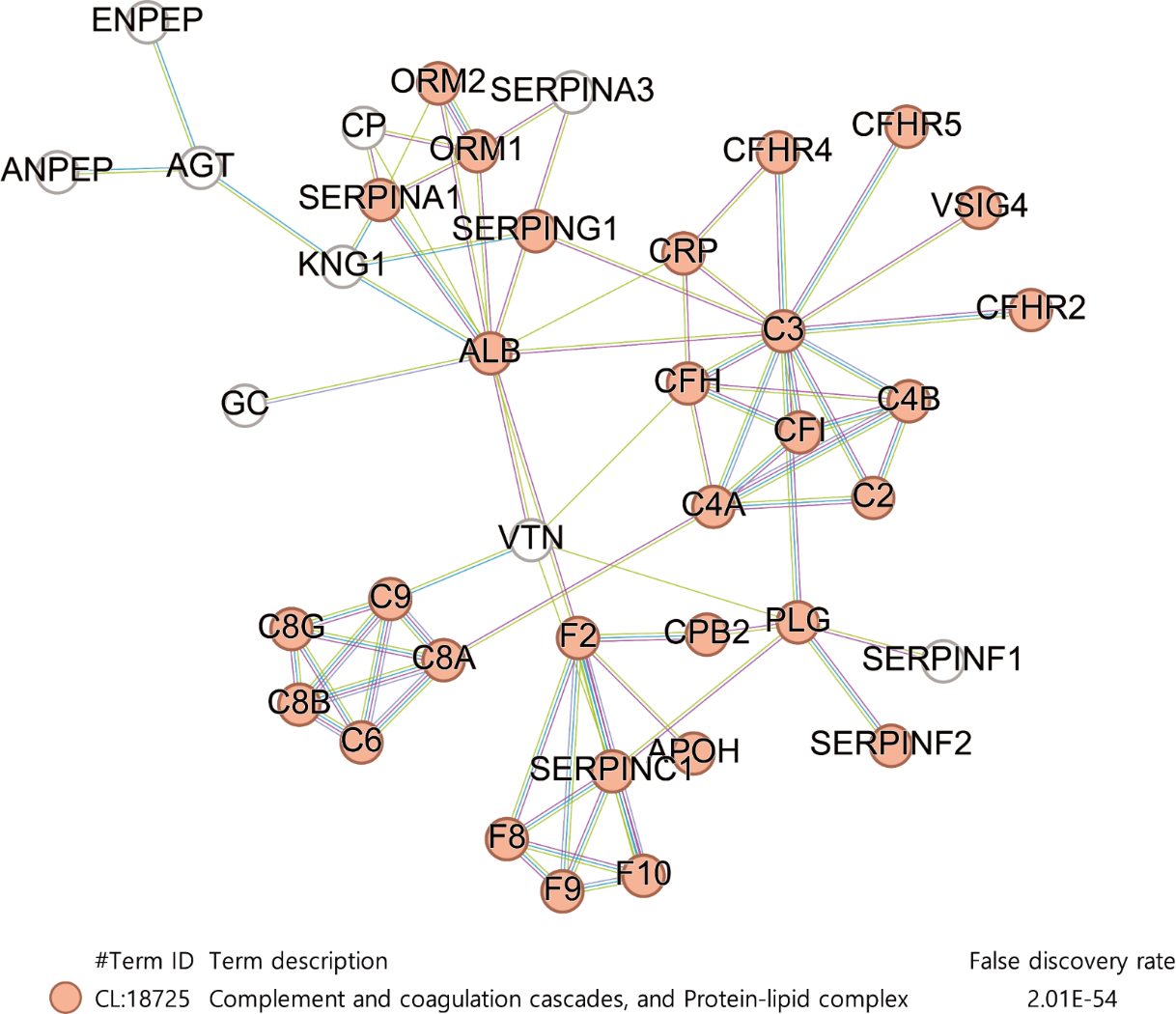

**Figure S10. STRING PPI analysis of the proteins in the M7 module.**

This STRING PPI analysis result contains 72 nodes with 75 edges. The minimum required confidence score was set to 0.9 (average node degree: 2.03, average local clustering coefficient: 0.409, and PPI enrichment P < 1.0e-16). The networks formed with four or fewer nodes were removed. Protein-protein interaction analysis was conducted using STRING based on the parameters of text mining (yellow edges), experimental evidence (purple edges), and database information (blue edges).

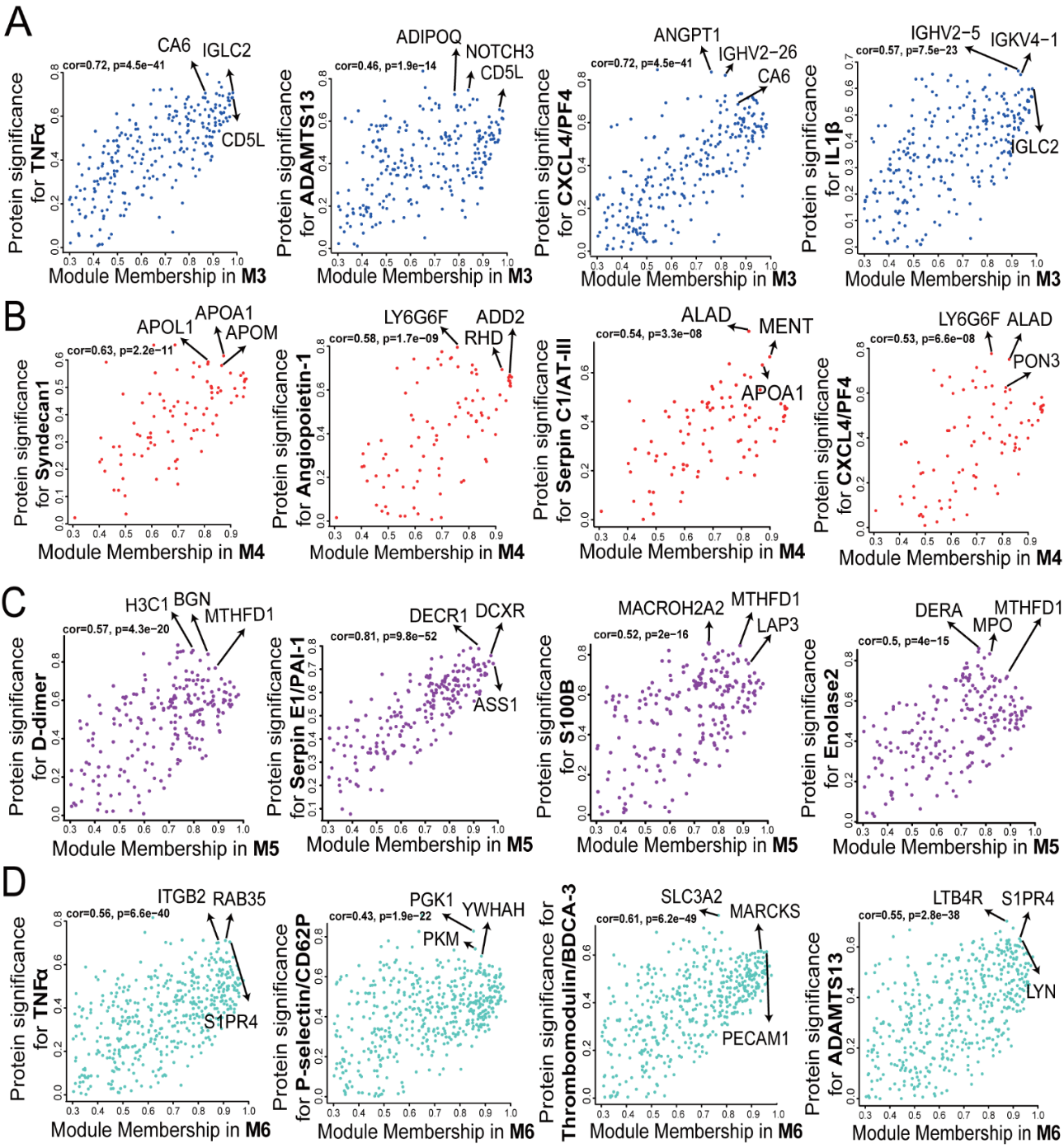

**Figure S11. The** **Module membership (MM) and protein significance (PS) plots in the M3-M6 modules.**

The traits in Luminex that correlate to M3 ~ M6 modules in *P* ≤ 0.01 were selected to analyze MM-PS plots and find significant proteins in sepsis pathophysiology. (A) MM-PS plots show the relationship between module membership (MM) for the M3 module and protein significance (PS) for the top 4 traits except diagnosis (TNFα, ADAMTS13, CXCL4/PFA4, IL1β). The key driving proteins were marked by black arrows. The protein list in the plot is presented in **Supplemental Table S8**. (B) MM-PS plots show the relationship between MM for the M4 module and PS for the top 4 traits except diagnosis (Syndecan1, Angiopoietin-1, Serpin C1/AT-III, CXCL4/PF4). (C) MM-PS plots show the relationship between MM for the M5 module and PS for the top 4 traits except diagnosis (D-dimer, SERPINE1, S100B, and Enolase2). (D) MM-PS plots show the relationship between MM for the M6 module and PS for the top 4 traits except diagnosis (TNFα, P-selectin/CD62P, thrombomodulin/BDCA3, ADAMTS13).
